# Moving out and moving on: the impact of mobility in a context of union dissolution on antidepressants intake in Belgium

**DOI:** 10.1101/2024.04.30.24306615

**Authors:** Joan Damiens, Christine Schnor, Didier Willaert

## Abstract

This research investigates the role of residential mobility in the relation between union dissolution and antidepressants intake. The dataset gathers information on 20 to 64-year-old individuals affiliated with the Belgian socialist health insurance fund – the largest public health insurance fund in French-speaking Belgium – and who lived in marital or non-marital opposite-sex partnerships in 2008 and separated between 2009 to 2018 (N=68,048). We used antidepressant consumption (>= 90 defined daily doses per year) as a dependent variable and conducted random-effect logistic regression models. Controlling for observed and unobserved individuals’ characteristics, we found that, mobility – defined by a change of municipality – during the year and/or the year following the separation is not associated with higher or lower antidepressants intake than staying on the previously shared place after the separation. However, we observe anticipatory effects for women: women who moved during their separation year had a higher medication use than women who stayed in the previously shared municipality. Repartnership is associated with lower antidepressants consumption for women, especially if they leave the shared place.

## INTRODUCTION

Recent health statistics from 2019 reveal that 7% of Europeans (EU-27) claim to suffer from chronic depression (Eurostat, 2021a). In Belgium, about 8% of the population reported to suffer from depression in 2018 (Gisle et al., 2018). Negative life events, such as union dissolutions, are associated with poor mental health outcomes (Bruce & Kim, 1992; Wyder et al., 2009). In parallel, the number of divorces per 1,000 marriages has risen over the last decades (Eurostat, 2021b) and the demand for housing increased as well, which makes it difficult for recently separated individuals and parents to find a decent and affordable place, especially in a context of post-separation economic loss (Biotteau et al., 2019; Feijten, 2005). To our knowledge, the role of residential mobility in the context of a union dissolution on mental health outcomes has been very rarely studied. This is surprising, as separation and mobility often go hand in hand: when a couple breaks up, at least one of the partners moves out of the joint home (Feijten & Van Ham, 2007). Residential mobility can bring both positive and negative outcomes on mental health: positive, as mobility can be a source of new opportunities and can help to move on from the past relationship (Mulder & Van Ham, 2005; Oishi, 2010; Trigg, 2009); and negative, as it can imply a break in close social ties, a fast and demanding change that can lead to and a loss of housing quality (Oishi, 2010), and a loss of life satisfaction (Oishi & Schimmack, 2010; Oishi & Talhelm, 2012; Stanley et al., 2012).

This article aims at investigating the antidepressants intake before, during and after a union dissolution, considering married as well as non-married cohabiting couples, and to evaluate how residential mobility at the time of separation impacts this relation. We draw data from members of the Belgian socialist health insurance fund, the largest health insurance fund in French-speaking Belgium, and the second largest on the national level (3 million members, out of more than 11 million Belgian residents). Our sample includes members who separated in the period from 2009 to 2018. Mental health is measured by looking at the yearly average consumption of antidepressants.

## BACKGROUND

### Residential mobility: both a challenge and a source of opportunities

According to the literature, residential mobility is associated with mixed consequences on mental health, depending on its context and the motivation (Choi & Oishi, 2020; Oishi & Schimmack, 2010). On the one hand, mobility is an important asset for individuals’ development. It can create new professional and personal opportunities, by giving more job possibilities (especially for men: Mulder and Van Ham, 2005), improving the interpersonal skills, and enlarging the social network (Oishi, 2010), as well as bringing one closer to non-resident family members (Mulder, 2018; Mulder and Wagner, 2010). Also, places carry symbols and memories, including traumatic elements (Trigg, 2009), and mobility can help move on from the past.

On the other hand, according to the *familiarity-liking theory*, moving and adapting to the new environment requests individuals to go out of their comfort zone, to change their habits and lifestyle (Magdol, 2002; Oishi & Talhelm, 2012). It can imply a break in social ties and a disruption in the feeling of belonging to a neighbourhood and being socially included. The damaging effect can be reduced if one disposes of high social capital that is not tied to the place of living, a high income and a mobility history (Mellor & Edelmann, 1988; Oishi, 2010; Stanley et al., 2012).

Residential mobility is a growing source of interest for public health and social sciences, but very little is known about how the context of a move can impact one’s mental health (Choi and Oishi, 2020). To our knowledge, there is very little empirical literature about the relation between residential relocation and mental health. Some previous articles highlighted the negative relation between mobility and physical good health. In a Dutch study, internal migrants reported more health issues than non-migrants (Verheij et al., 1998). In an Australian study, middle-aged women who relocated within the country presented a higher risk of chronic illness than their counterpart who did not move, and both men and women had more contact with a specialised doctor when they were internal migrants (Larson et al., 2004). But when it comes to psychological implications of mobility, very little is known. Studies on home eviction could show the detrimental impact of this forced move on adults’ and children’s mental health (Vásquez-Vera et al., 2017). A qualitative study conducted in Dusseldorf could show that migrants were in general less happy than locals (Hendriks et al., 2016). This result is interpreted based on the daily life activities: migrants tend to be less integrated in a social network, and thus less involved in activities that promote wellbeing, health and mental health (Hendriks et al., 2016). A qualitative Chinese investigation could confirm this conclusion and the idea that internally mobile individuals are likelier to feel isolated and unsatisfied by their close social environment (Liu et al., 2017). A recent qualitative Australian research (Wood et al., 2023) recalled the lack of research on this relation and could put forward that the residential move’s association with mental health was highly related to the financial context of the move, the ability to afford the housing, the load of the credits as well as the tenure transitions: moving to accede homeownership was associated with positive mental health outcomes, unlike moving in a context of homeownership loss (Wood et al., 2023).

### Union dissolution and mental health

Residential mobility is closely related to partnership changes, and particularly the physical separation of a couple. After a separation, in most cases, at least one of the two ex-partners will quickly move out of the family home. Being separated or divorced is associated with an increased risk of residential mobility, compared to married or single individuals, for both men and women (Kulu et al., 2021): this higher risk peaks at exact moment of separation, and decreases over time. In some countries, such as Belgium, the probability of a move remains higher for separated individuals for a longer time – more than a year – after the union dissolution (Kulu et al., 2021).

There is already an extensive theoretical and empirical literature on the negative relation between union dissolutions and mental health. In the early 1990s, psychiatric theories explained that a separation is often accompanied with an increase in psychological distress (Booth & Amato, 1991). Empirical studies could confirm that the levels of mental health started to decrease before a marital or non-marital union dissolution, as shown by longitudinal studies in several contexts (US: Rhoades et al., 2011; Sweden: Switek and Easterlin, 2018; Norway: Næss et al., 2015), and with several measures of mental health (in terms of antipsychotics consumption rates: Metsä-Simola and Martikainen, 2014; in terms of depression: Tosi and van den Broek, 2020). A study conducted in New Haven (US) confirmed that marital disruption was associated with an increased risk of a first depressive episode (Bruce & Kim, 1992).

A union dissolution can rather be considered as a process than as an event: separation can take several months or even years and is preceded and followed by negotiations and legal aspects (Schnor, 2015). The investigation of the relation between separation and mental health must then account for the periods before, during and after the separation: the anticipation to this stressful event and the conflictual environment can change individual’s behaviours (e.g. women starting to work (again) before a divorce: Vignoli et al., 2018) and trigger a decline in mental health (Næss et al., 2015; Rhoades et al., 2011; Switek & Easterlin, 2018).

A separation is synonym of short-term and long-term changes in one’s life and habits, which can impact wellbeing and life satisfaction. Among them, we can cite the loss of resources, the new configuration of the family links – including the custody of children, the relation with the in-laws and the common friends, as well as the change of residence and living conditions. The factors that explain the loss of wellbeing after a union dissolution are shown to differ between men and women. Men are more vulnerable for short-term consequences of union dissolutions – such as the loss of social support – than women (Leopold, 2018). On the longer term, women have to face a higher risk of poverty and single motherhood, which can have multiple consequences on their lives and be detrimental for their mental health, for a longer time (Conejero et al., 2018; Fernquist & Cutright, 1998; Leopold, 2018; Stack, 1982). Low partnership quality and the threat of separation can lead to an anticipation effect (a decrease in wellbeing that starts before the separation is enacted) and some long-term effects that can years after the relation ended. Long-term effects are likely to be gendered, as women suffer more from long-term consequences than men (Leopold, 2018). Entering a new cohabiting union can be associated with a socioeconomic improvement for divorce and separated individuals, especially for women (Dewilde & Uunk, 2008).

Belgium presents higher divorce rates than the EU average since the early 1980s. For the year 2019, the country counted 50.7 divorces per 100 marriages (Eurostat, 2021b). But contrary to other European countries, the number of divorces tended to decrease in Belgium over the last decades (Eurostat, 2021b). This also reflects a higher selectivity of marital unions, that is accompanied by an increase in non-marital cohabitation and age at first marriage (Statistics Belgium, 2020). The dissolution of non-marital cohabitations is not registered in official statistics, but survey data suggests that cohabitations are less stable than marriages (Andersson et al., 2017).

### Mobility in a context of union dissolution

Residential relocation is very little studied as a determinant of the mental health struggles post-separation. Nonetheless, among all the difficulties and psychological struggles that can happen during a union dissolution, the change of housing is worth study. First, the context of a post-separation move is far from ideal. The new residence is often found in a context of emergency and fewer resources and might be a temporary option (Feijten & Van Ham, 2007), such as moving in (at least temporarily) with a friend, a parent or a sibling (Feijten & Van Ham, 2007).

Individuals can expect to lose some housing quality and homeownership status (Lersch & Vidal, 2014), compared to other types of move. Second, poor quality and instable housing are detrimental for mental health, as it questions one’s trust in the future, self-esteem and social relations (Bernard, 1998; Evans et al., 2003; Magdol, 2002; McCormick et al., 2012; Oishi & Schimmack, 2010; Shklovski et al., 2006; Stokols et al., 1983). Third, due to the possibly conflictual climate that surrounds a union dissolution and the redefinition of the family unit, separation-driven moves can damage the social network of a person and trigger social isolation (Heikkinen et al., 1993): for parents, moving can be detrimental to the relation with the children, especially for the parent who has no custody of the minor children or a custody arrangement that does not encourage their relation with their children (Ferrari et al., 2019). The moves after a separation are characterised by a high instability, visible through repeated episodes of mobility, which can be expected to amplify the detrimental impact of this life event on mental health. Several moves can indicate new opportunities, but also that the individual faces economic hardship that does not allow him or her to afford their housing alone, that the immediate new place of living might be inadequate for their needs, or that their overall situation – regarding the custody of the children, for instance – is uncertain (Feijten & Van Ham, 2007).

Right after separation, the decision of who moves and who stays in the previously shared place is a complex information that can rely on economic and contextual factors. In Belgium, the legal framework does not encourage any party to leave or stay in the shared place. According to a *cost-benefit approach*, we can assume that if one does not have enough financial means to afford those costs, he/she will likely move (Mulder & Malmberg, 2011; Mulder & Wagner, 2010). Studies for Belgium confirmed women’s higher risk to move after their separation, especially among the lower-educated ex-couples (Theunis et al., 2018). Beyond that, the decision of who moves and who stays in the previously shared place masks the decision process of the separation itself. Even though there are no inflexible rules, we can assume that the person who decides to break up is likely to be the person who moves out of the shared place, because they are more ready to leave the relationship or because they plan to join a new partner (Kolodziej-Zaleska & Przybyla-Basista, 2020; Symoens et al., 2013). Women tend to initiate more often the separations than men (Hewitt et al., 2006). Among those contextual elements, the reconstruction of a new partnership in the two years after the separation matters in the relation between the mobility and mental health, as it can indicate that the individual in a new relationship is ready to move on from the past relationship. In this specific context, mobility appears as the first step of a new life chapter and a new union formation.

### Measuring mental health

Some qualitative studies could put forward the negative association between internal migration and wellbeing, but to our knowledge, their definition of wellbeing was mainly self-reported and based on an appreciation of happiness and life satisfaction. On the opposite, research on the relation between separation and wellbeing, very diverse ways to measure mental health are suggested, such as depressive symptoms (Tosi & van den Broek, 2020), antidepressants consumption (Metsä-Simola & Martikainen, 2014) and suicidal behaviours and mortality (Bruce & Kim, 1992).

The use of antidepressants to measure depression is commonly used in studies that focuses on mental health (Eriksson et al., 2004; Frandsen et al., 2016). Such an indicator represents both positive and negative aspects. On the one hand, it gives a quantifiable and objective measurement of mental health struggles and a depressive state. In Belgium, antidepressants can be prescribed mainly by general practitioners, psychiatrists, or any other specialised doctor, to treat mainly depressive states, but also, anxiety, burn-out, migraines and some muscular pain conditions (Boutsen et al., 2015; Cascade et al., 2007). One condition to be diagnosed as depressive is to encounter depressive symptoms for at least two continuous weeks, which makes a difference between sadness and depression (*Depression*, s. d.). This treatment is totally reimbursed by the social insurances, which is compulsory and affordable for a very large part of the population living in Belgium.

On the other hand, there is a risk of underestimation of depression with this measurement. Antidepressants prescription requires a professional diagnostic but also some treatment adherence (Anderson & Roy, 2013). The use of antidepressants as a predictor for depression can thus be interpreted as the lower bound of depression prevalence. It has been shown to underestimate depression among certain populations, especially in the case of non-adherence to the treatments (Fried & Nesse, 2015; Uher et al., 2012).

Studies show that antidepressant consumption varies according to individuals’ characteristics, independently from their depressive symptoms. A report from by the socialist health insurance fund showed that in 2012, 27% of the suicide attempt survivors who were hospitalised had not contacted a doctor or psychiatrist during the last three months before their attempt (Boutsen et al., 2015). Seeking for mental health care de pends on many factors. Men tend to open less about their mental health issues and seek less medical help for them (Payne et al., 2008; Van der Heyden et al., 2009). Within Belgium, lower social strata tend to consume more antidepressants than higher social strata (Gisle et al., 2018), as the latter have more financial resources to rely on other solutions such as psychotherapies (Hilvert-Bruce et al., 2012). Personal beliefs about mental health medication and cultural background, as well as social relations – such as the antidepressant consumption of the close family and friends – can also impact individual adherence to a treatment (Anderson & Roy, 2013; Chakraborty et al., 2009; Garfield et al., 2004; Ho et al., 2017), especially in a context of controversy about mental health medication effectiveness and side effects (Healy & Whitaker, 2003; Isacsson et al., 2005).

### RESEARCH QUESTIONS AND HYPOTHESES

Some qualitative studies could put forward the negative association between internal migration and life satisfaction, but to our knowledge, no previous study investigated how residential mobility can interfere in the relation between union dissolution and antidepressants treatment. The objective of this paper is to discuss how residential mobility can be associated with antidepressants intake increase during a union dissolution. There is very little research on the relation between mobility and mental health, but theoretical work highlighted the importance of the context of the move on its implications on life satisfaction and wellbeing. On the one hand, mobility in a context of separation is a specific type of mobility. It is associated with a negative life event, with emergency, with a decrease of resources at the level of the household and with a reconfiguration of the family ties and relations. The cumulation of two negative events, namely the breakup and the subsequent residential mobility, would make these demanding life changes even more intense, creating more distance with the family members and friends, requesting fast life adaptations**. In a first hypothesis, we assume that individuals who move during the year of the separation would present a higher antidepressants consumption than individuals who stay in the previously shared place.**

In addition, men and women face different consequences after a separation. Men often struggle with short-term obstacles related to loss of social support, while women tend to suffer longer, from social and economic difficulties (Ferrari et al., 2019; Leopold, 2018). Also, women tend to move out of the previously shared place more often, to have more instable housing paths after a separation (Ferrari et al., 2019), and to initiate more the separation (Hewitt et al., 2006). **As a second hypothesis, we expect gender-related specificities in the results. A) Because they are likelier to initiate the separation and to anticipate the imminent union dissolution and subsequent move and life changes, women who will leave the housing would present a higher antidepressants consumption before the separation than women who stay in the previously shared place. B) After the separation, men would show a higher antidepressants intake when they move than when they stay, as moving would trigger a higher risk of isolation and loss of social support. C) For women, the gap of antidepressants intake between those who move and those who stay would be less marked: women are associated with more economic losses after a separation and staying in the previously shared place means they keep carrying the financial load of this housing on their own, with more limited resources.**

The possibility to enter a new relationship can change the observed relation. First, a post-separation move could result from the project to end a cohabitation and to start a new relationship. In addition, repartnerhsip tends to protect individuals, especially women, from socioeconomic loss. **As a third hypothesis, we assume that moving in a context of union dissolution is associated with a lower antidepressants consumption when the individual repartners in the following years, than when they move to stay single. We assume this advantage of repartnership to be even more visible for women, who would be more protected from post-separation socioeconomic losses if they live with a new partner.**

## Data

### DATA AND METHODS

To answer those questions, it is possible to rely on health insurance data for medical information, which can be merged with National register that provides information about the partnership transitions and the residential relocations. More specifically, we use population panel data on individuals living in Belgium and affiliated to the Socialist Health Insurance Fund (Solidaris, SHIF)^1^. Our sample includes marital and non-marital couples who were intact on January 1^st^ 2009 and who went through a union dissolution between 2009 and 2018: 33,101 men and 34,947 women^2^ (Table 1). It is important to note that we only considered couples formed by two opposite-sex partners who were living together and who are both affiliated to the SHIF during the whole period 2009-2018, to get information about the partner. Note that this condition is also necessary to be able to identify couples and eventually their separation. To keep the sample balanced, we excluded couples in which one member left the observation before 2018, by moving abroad, dying or changing health insurances. Finally, we restricted our observation to the population aged 20 to 54 in 2009 – and who will turn 30 to 64 at the end of the observation period, as the intake of antidepressants among younger or older persons is particularly influenced by specific factors. For instance, the use of antidepressants among children and teenagers is controversial in psychiatric studies and practice, as the determinants of mental health for young people are even more diverse than for adults (Korenblum, 2004; Morrison & Schwartz, 2014). Antidepressants are often prescribed to elderly individuals as well, especially in case of mobility loss or cognitive diseases (Doraiswamy et al., 2003; Karkare et al., 2011).

**Table 1.** Sample characteristics of the analytical sample (intact heterosexual couples in 2009 who will separate in 2009-2018, formed by two members of the SHIF), at the beginning of the observation period (01/01/2009) and according to mobility status during the separation year, in numbers and percentage.

| **a/ MEN** |  |  |  |
| --- | --- | --- | --- |
|  | **a)** | **b)** | **c)** |
|  | **TOTAL** | *Moved at t* | *No move at t* |
|  | **33,101** |  |  |
| **Total** |  | 9,345<br>28.23% | 23,756<br>71.77% |
| Living in a marital union | 16,745<br>50.59% | 4,699<br>50.28% | 12,046<br>50.71% |
| Living in a non-marital union | 13,473<br>40.70% | 3,815<br>40.82% | 2,658<br>80.18% |
| Living with children | 7,725<br>23.34% | 2,193<br>23.47% | 5,532<br>23.29% |
| Childless or not living with children | 22,493<br>67.95% | 6,321<br>67.64% | 16,172<br>68.08% |
| Flanders | 14,890<br>44.98% | 3,953<br>42.30% | 10,937<br>43.20% |
| Wallonia | 14,956<br>45.18% | 4,100<br>43.87% | 10,856<br>47.90% |
| Brussels | 3,255<br>9.83% | 1,292<br>13.83% | 1,963<br>8.26% |
| Unemployment | 4,135<br>12.49% | 1,148<br>12.28% | 2,987<br>12.57% |
| Incapacity | 379<br>1.14% | 88<br>0.94% | 291<br>1.22% |
| Increased reimbursement | 3,085<br>9.32% | 883<br>9.45% | 2,202<br>9.27% |

| **b/ WOMEN** |  |  |  |
| --- | --- | --- | --- |
|  | **(a)** | **(b)** | **©** |
|  | **TOTAL** | *Moved at t* | *No move at t* |
|  | **34,947** |  |  |
| **Total** |  | 9,991<br>28.59% | 24,956<br>71.41% |
| Living in a marital union | 18,186<br>52.04% | 4,728<br>47.32% | 13,458<br>53.93% |
| Living in a non-marital union | 13,776<br>39.42% | 4,362<br>43.66% | 9,414<br>37.72% |
| Living with children | 7,991<br>22.87% | 2,783<br>27.86% | 5,208<br>20.87% |
| Childless or not living with children | 23,971<br>68.59% | 6,307<br>63.13% | 17,664<br>70.78% |
| Flanders | 15,805<br>45.23% | 4,430<br>44.34% | 11,375<br>45.58% |
| Wallonia | 15,692<br>44.90% | 4,549<br>45.53% | 11,143<br>44.65% |
| Brussels | 3,450<br>9.87% | 1,012<br>10.13% | 2,438<br>9.77% |
| Unemployment | 4,911<br>14.05% | 1,312<br>13.13% | 3,599<br>14.42% |
| Incapacity | 528<br>1.51% | 156<br>1.56% | 372<br>1.49% |
| Increased reimbursement | 3,779<br>10.81% | 859<br>8.60% | 2,920<br>11.70% |

Source: Belgian socialist health insurance fund.

Using civil status information and household configuration from the National Register, we can directly identify marital couples. For non-marital couples, we relied on several assumptions, such as having only two opposite-sex adults aged 16 and above living in the same household, with an age difference lower than 15 years between them. These assumptions exclude same-sex couples and couples with a large age gap, but their consistency is supported by external validation: the applied definition captures more than 90% of the non-married partnerships that were self-declared in the Generations and Gender survey conducted in 2009 in Belgium (Lodewijckx & Deboosere, 2011).

This dataset is particularly adapted to answer our questions, as they give us the yearly consumption of antidepressants in defined daily doses, they cover individuals in the years before, during and after a union dissolution and follow their intermunicipal relocations. However, they present some limitations that need to be addressed, but that do not prevent this dataset to be adapted for our objectives.

First, the information is yearly, which does not allow us to reconstruct precisely the trajectories of the individuals, like the number of moves or separations during one year. However, this does not fragilise our methodological framework, and we assume that these very fast repartnerships that do not last a year are quite rare.

Second, we cannot measure intramunicipal moves, only changes in municipalities. This implies that we do not capture all moves, particularly the very short-distance ones, that are particularly common for parents sharing custody (Dewilde & Uunk, 2008). However, Belgium counts a bit less than 600 municipalities, with an average of 50 km² of area each. In such a context, we can assume that changes of residence without a change of municipality do not imply a strong change in life habits and social links close to home. Our dataset gives us the information we need in terms of residential mobility, that is a change in environment.

Third, the population covered by the SHIF cannot be considered as representative of the whole Belgian population. In Belgium, the health system is based on the obligation for each resident to adhere to a health insurance fund, to cover at least partially the health-related costs, in terms of therapeutic or diagnostic actions, medical interventions, hospitalisations or treatments (D’hoore & Stordeur, 2004). Historical, socioeconomic and cultural factors led to the creation of several social movements, such as trade union, youth movements or religious communities. Health insurances are the continuity of these different movements and are divided in three large pillars: Christian, socialist and liberal health insurances (Faniel & Gobin, 1992). Individuals also have free choice between several health insurance companies, whose basic fees are comparable. According to their position in society, to their beliefs or values, to their family history, individuals will feel closer to a certain health insurance. This means that every health insurance will cover a different population, with different demographic and socioeconomic characteristics. The SHIF covers less than a third of the Belgian population (about 3 200 000 members in 2018, i.e. 28% of the population), with a large representation of the Walloon population, living in the southern French-speaking region of the country (1 300 000 Walloon members in 2018, i.e. 38% of the Walloon population).

### How to measure mental health?

Our data includes every individual’s yearly antidepressants prescription, in Defined Daily Doses (DDD) which is the assumed average maintenance dose per day for a drug used for adults (Sinnott et al., 2016). With this information, we want to differentiate a short-term prescription of antidepressants to diagnosed depression, which requires a treatment of at least 3 months. A 90 DDD threshold is shown to be the adequate minimum duration of treatment of depression, in order to feel better and avoid relapses (Hirschfeld, 2001; Moustgaard et al., 2014). Our outcome variable is thus dichotomous, distinguishing yearly consumption of 0-89 DDD and 90 DDD and more. In robustness checks, we also tested variation of the antidepressant consumption in DDD, without any threshold.

### How to measure separation?

We measure separation by looking at the household identifiers. We assume that, between years t and t+1, if the two partners still have the same household identifier, nothing changes in the relationship. On the contrary, if at least one of the partner changes household identifiers and if the two partners have different household identifiers, this means the couple does not share the same accommodation any longer, which defines a union dissolution. As one of our objectives is to investigate the timing of union dissolution, a categorical variable distinguishes: 1) two years and more before the separation; 2) the year before the separation; 3) the year of the separation; 4) the year following the separation year; and 5) two years and more after the separation. Here, we can only consider the first separation of the individual in the observation period. The household composition variable allows us to estimate the civil status and the parental status of the individual, but after this first relationship, we do not have any information about the characteristics of the possible new partner if they are affiliated with another insurance. Using this variable, we estimate that about 24.6% of men and 22.3% of women from our sample will repartner before the end of our observation period.

### The moderator effect of mobility

Our second objective is to examine whether the effect of separation on mental health is moderated by residential mobility. The definition of a residential move is a change of the municipality of residence between year t and t+1, recoded in a dichotomous variable. The relation between separation and mental health according to the mobility status during the separation year is studied by using different approaches. A) First, in a time-varying approach, the individual who relocates during the separation year is assigned to the “mover” category only from the moment of the separation; B) Second, we take an anticipatory time-constant approach, in which people who will move during their separation year is assigned to the “mover” category throughout the whole observation period – both before and after the separation – to evaluate possible anticipation effects: we question the possibility that a future (and anticipated) mobility could increase antidepressants before the separation; C) we investigate the role of repartnership in the relation, by distinguishing 4 trajectories after the separation: i) individuals who did not move during separation year (t) and who do not repartner; ii) individuals who did not move in year t but repartnered; iii) individuals who moved in year t and did not repartner; iv) individuals who moved in year t and repartnered. This allows to know more about the context of the move and study the moves that can be attributed to possible new union formations.

### Control covariates

The database includes a range of characteristics for each individual, gathered on January 1st of each year. First, we distinguish men and women, as mental health problems, medication consumption and effects of separation on (mental) health differ by gender (Payne et al., 2008; Van der Heyden et al., 2009). We treat age as a continuous variable. We also include geographic information in terms of region of residence: Flanders, Brussels-Capital Region and Wallonia. The regions differ in terms of mental health policies, with regional budgets and authorities managing mental healthcare. Then, the household type variable is based on the composition of the household (Van Imhoff & Keilman, 1992), and allows to estimate the marital and parental status of the individual, which matter for mental health (Graham, 2015; Rhoades et al., 2011).

The available socioeconomic information is also included. We know that a lower socioeconomic category is associated with a higher risk of mental health issues and tend to decrease the likelihood to take up therapy without medication (Ahmad & Ismail, 1988; Wei et al., 2005). We estimate socioeconomic precariousness with two variables. When the household’s income is low enough or based on social benefits, they are automatically entitled to an increased reimbursement of the healthcare: a dichotomous variable indicates whether the individual benefits from this increased reimbursement. The access to this status does not depend on the healthcare expenses and is attributed automatically, based on the declared income of the household. As it is known that unemployment is closely related to mental health (Artazcoz et al., 2004), we also used information on the number of days worked over the year as a proxy for employment status. One is considered unemployed and receiving unemployment benefits if the number of days worked is below 150 days per the year (less than 6 months). We accounted for the possibility of this 6 months period to overlap two calendar years.

Moreover, studies put forward that physical and psychological pain are very interrelated (Conejero et al., 2018). The general health status is measured by counting the number of days that an employee received sickness or invalidity benefit. We consider individuals in a disability state if their number of days with sickness or invalidity benefit exceed 150 working days a year.

We also account for the antidepressant consumption of the (ex-)partner, as it can influence one’s openness to mental health medication and adherence to the treatment (Dupre and Meadows, 2007^3^). As both the members of the couple are affiliated to the SHIF, this information is directly available. We account for changes in main antidepressants prescriber between two consecutive years. This is important as a residential move may lead to a change in medical doctor(s). The patient may be less open about his/her mental health problems to a new healthcare profession or may get a new diagnosis, which can impact the treatment. Finally, the distance of the move was added to the models, through a categorical variable, distinguishing whether the distance between the centroids of departure and arrival municipalities centres were located 16 kilometres away for each other at the most, or above this threshold. In Belgium, we can consider than 16 km is the threshold to define long-distance moves, as 16 km is the radius of the largest agglomeration in terms of area (Tournai). All those variables are time-varying, but some of them are likelier to vary in time (unemployment days, marital status, region of residence and parental status to a lesser extent) than others (sex does not vary, increased reimbursement, low variation of prescribers).

## Methods

The variation of antidepressants intake might depend on observed and unobserved characteristics of the individuals, such as mental illnesses, genetic predispositions, or other health issues. To estimate the depression risk in a context of panel data, we chose to run random-effect logistic models that are presented as the logarithm of the odds of depression:

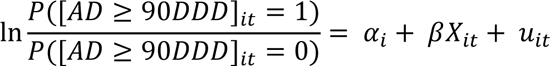

Where *i* is each individual (from 1 to N), *t* is time in years, α_*i*_ is the individual specific effect, β_*it*_ is a vector of parameters to be estimated, *X*_*it*_ is a vector of explanatory variables (including union dissolution, residential mobility and the interaction of both), and *u*_*it*_is the disturbance/error term following a normal distribution. We estimate logistic regression models, as the risk of the yearly antidepressants intake being 90DDD and above (0/1) is a binary variable. α_*i*_ is not directly observable. It can be considered as constant parameters (with fixed-effect logistic regression models) and as random parameters that are allowed to vary (with random-effect logistic regression models). We decided to model random-effect regression models, as fixed-effects models do not give estimates for time-constant variables (Bell et al., 2019; Halaby, 2004). Above that, we cannot assume that individual-specific effects are correlated with the predictors (predictors cannot capture the whole within-effect).

Interaction results will be presented in predicted probabilities of depression:

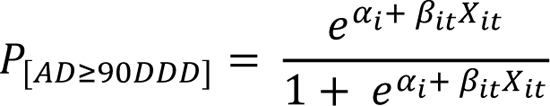

for each vector *X*_*it*_ of explanatory variables (including union dissolution, residential mobility and the interaction of both).

We conducted robustness checks to test other models’ specifications. First, linear regression models based on Ordinary Least Squares (OLS) were conducted by using, instead of the depression risk, the number of DDD of antidepressants consumed over a year. These model results are presented in Figures A1-A2 and display the predicted number of antidepressants DDD consumed during the separation steps and according to the mobility status at the time of the separation. Finally, Poisson regression models were estimated (Figures A3-A4), as antidepressants intake remain a rare event (Cai et al., 2010). Results of the Poisson models are expressed in incidence rates (in our case the incidence rates of depression for the populations who separates and moves, and for the population who separates and does not move). First, we reproduced the random-effect logistic regression models only on couples in which partners had not consumed any antidepressant in 2008, right before the observation period (Figure A5). This subsample counts 38,199 men and 40,161 women, i.e. 76% of the analytical sample. The objective of this check is to control whether previous mental health medication intake and predisposition to chronic depression may drive our results.

## Descriptive part

### ANALYSIS

In 2009, 12% of the socialist health insurance fund affiliated population consumed at least 1 DDD of antidepressants, and 7% of the population had an antidepressant intake of 90 DDD and more (Figure 1). This percentage increased over the observation period while remaining higher for men (8% in 2018) than for women (14% in 2018). The difference between men and women is particularly visible when comparing their couple trajectory from 2009 to 2018. Less than 10% of men who separated or divorced during the observation period consumed more than 90 DDD of antidepressants in 2018, compared to 19% of the women in the same situation.

**Figure 1.**
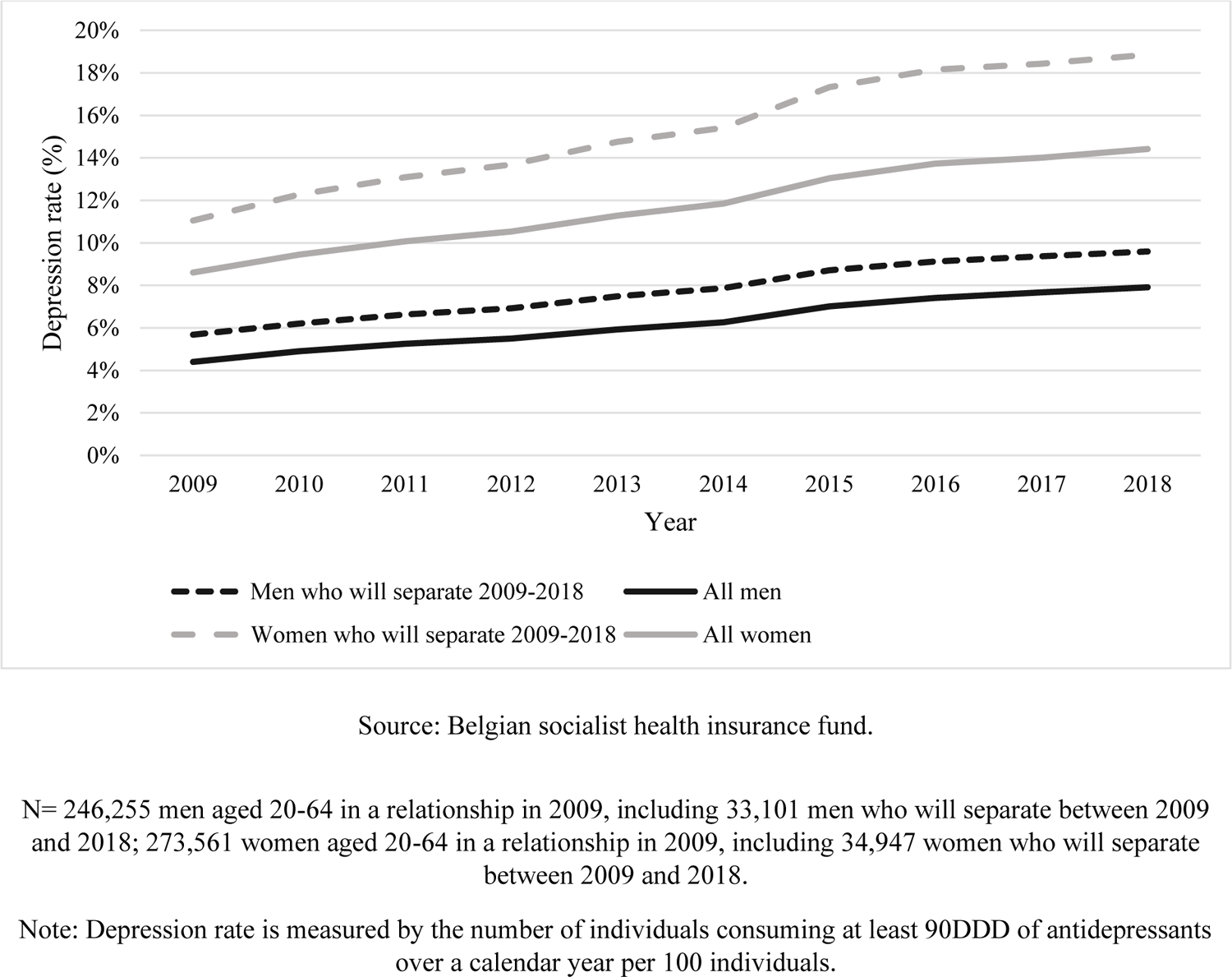
Rate of men and women whose antidepressants intake is at least 90DDD per year, year 2009 to year 2018.

Our sample includes 33,101 men and 34,947 women who went through a separation in the observation period and are followed over 9 years, resulting in 612,432 person-years. Table shows the characteristics of the sample. All our variables are time-varying, but some of them are more likely to vary (unemployment) than others (parental status). We notice that most of our sample was in a marital union before separating. Most of them also have children. A large part of our sample lives in Wallonia at the beginning of the observation period. We also estimate that 12 to 14% of our sample is unemployed in 2009, and a very small percentage cannot work at that time. About 10% of our sample has access to increased reimbursement due to low income. There are very few differences in characteristics at the beginning of the observation period between people who will move during the separation year b) and people who will not move or move but not change municipalities of residence c).

Other time-varying covariates will also be added in the models. One of them is the change of prescribers between two consecutive years. It can be estimated that among the moves in 2009, about 5% of the men and 9.1% of the women will change prescribers. Also, we add the distance of the move. In 2010 and 2015, 32% of the moves between municipalities will cover a distance of at least 16 km. These numbers are consistent over the years.

### Analytical part

All analyses are conducted on our main sample: individuals aged 20 to 54 in 2009, and who were part of an intact heterosexual couple in 2009 who will split up in 2009-2018, and that formed by two members of the SHIF.

Table 2 presents the results of random-effect logistic regression models in odds ratio of antidepressants intake being 90DDD and more, for men and women. Model 1 includes the separation timing indicator (2 years and more before, 1 year before, year of separation, 1 year after, 2 years and more after). Model 2 includes both this separation indicator and the mobility status during the separation year. Model 1 shows a clear increase in depression risk (antidepressant consumption) during the year before the separation, compared to the previous years. This anticipation effect of the separation is visible for both men and women, and after controlling for observable and unobservable individual characteristics. The antidepressants consumption peaks during the year of the separation and decreases after the separation, especially for men who reach the baseline depression level. For women, even two years and more after the separation, the antidepressants intake remains higher than it was two years and more before the separation. Model 2 includes both separation and mobility indicators. The odds of antidepressants intake being 90DDD and more over separation steps remains identical even after controlling for the moves during the separation year. There is no clear association between a move during the separation year and the medication use for men and women.

**Table 2.**
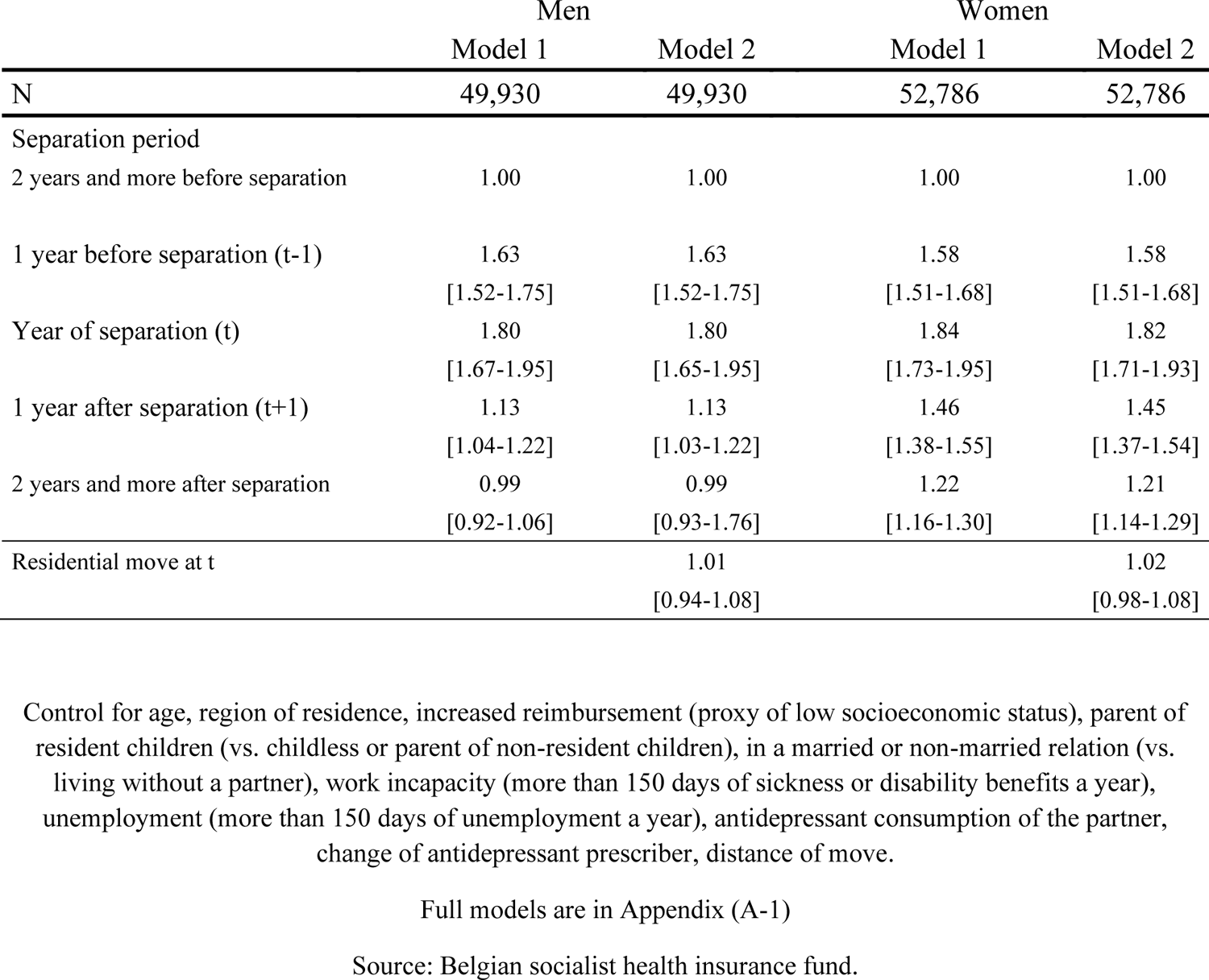
Random-effect logistic regression on the risk of having a antidepressants intake over 90 DDD (0/1) for men and women, expressed in odds ratio.

### First hypothesis

In Figures 2 to 5, we investigate the variation of the predicted probability of an antidepressants intake of 90 DDD and above, before, during and after union dissolution, according to men’s and women’s mobility status in the year after the separation.

**Figure 2.**
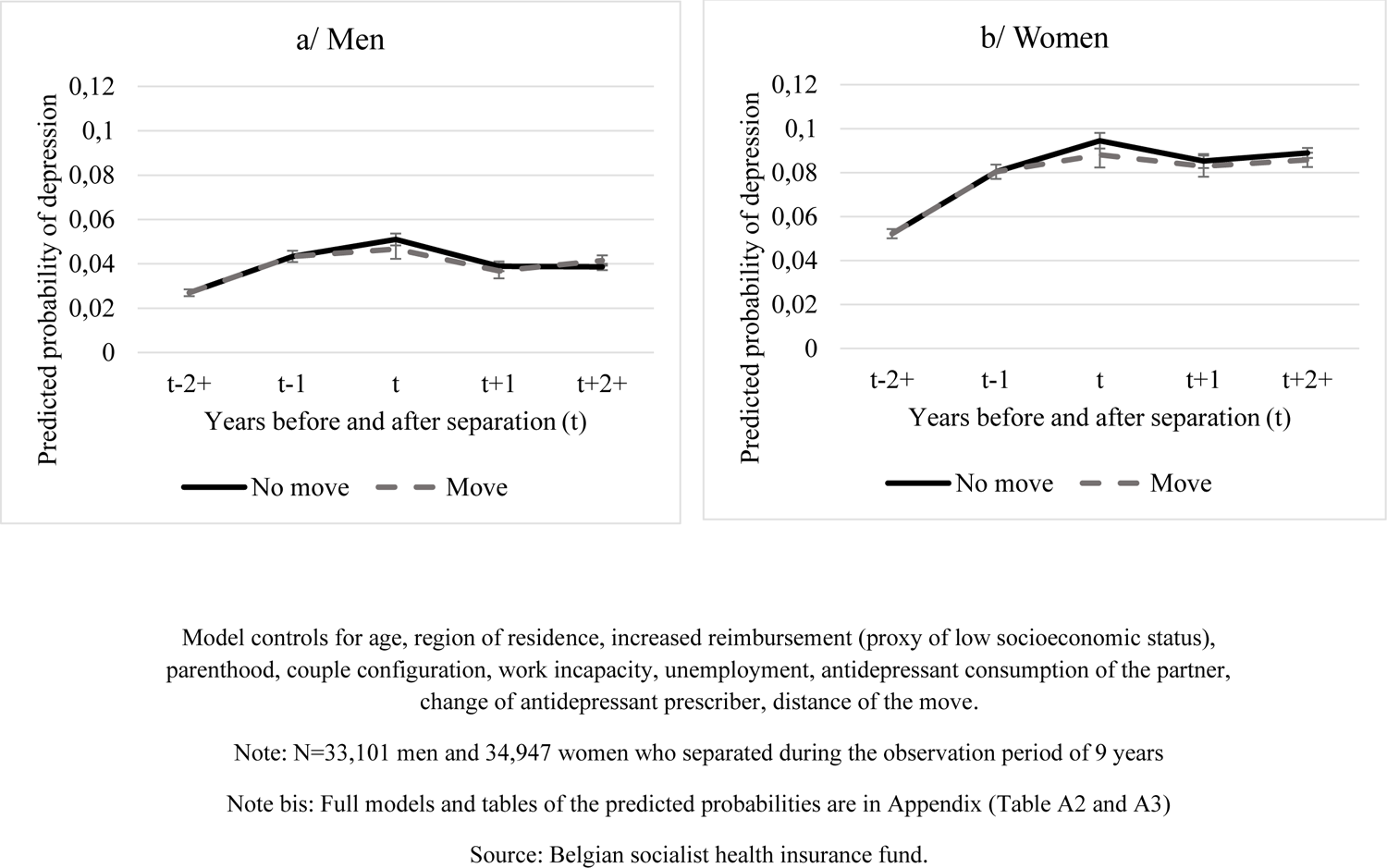
Predicted probability of antidepressants intake being 90DDD+ a year (based on random-effect logistic regression models) according to the mobility status of the individual at the time of separation (t). The mobility status during the year of separation (t) is attributed during this separation year (not before).

In Figure 2, we consider the mobility status as time-varying and assign individuals to the “move” category from the year of the separation and after if they moved in the separation year. After the separation year, men and women who did not make an intermunicipal move during the separation year present a slightly higher risk of antidepressants treatment during this specific year. For the following years, no differences are to be noted. All these results present a very low effect strengths, with overlapping confidence intervals, which does not allow us to conclude on our first hypothesis.

### Second hypothesis

We then conducted anticipatory analyses and considered the mobility status during the year of the separation as time-constant. In Figure 3, we assign the mobility status in the separation year for the whole observation period. Women who moved during the separation year show a higher risk of antidepressants treatment the year before the separation and then a convergence in terms of depression risk from the year of the separation. This goes in the direction of our second hypothesis and confirms the gender-specificity of the relation. We can confirm that women who will move anticipate the separation and present an increased antidepressants intake in the years before. After the separation, moving or staying in the previously shared residence does show much difference for women, nor for men. We expected men who moved to present a higher antidepressants intake than those who stayed, but our results do not confirm this idea.

**Figure 3.**
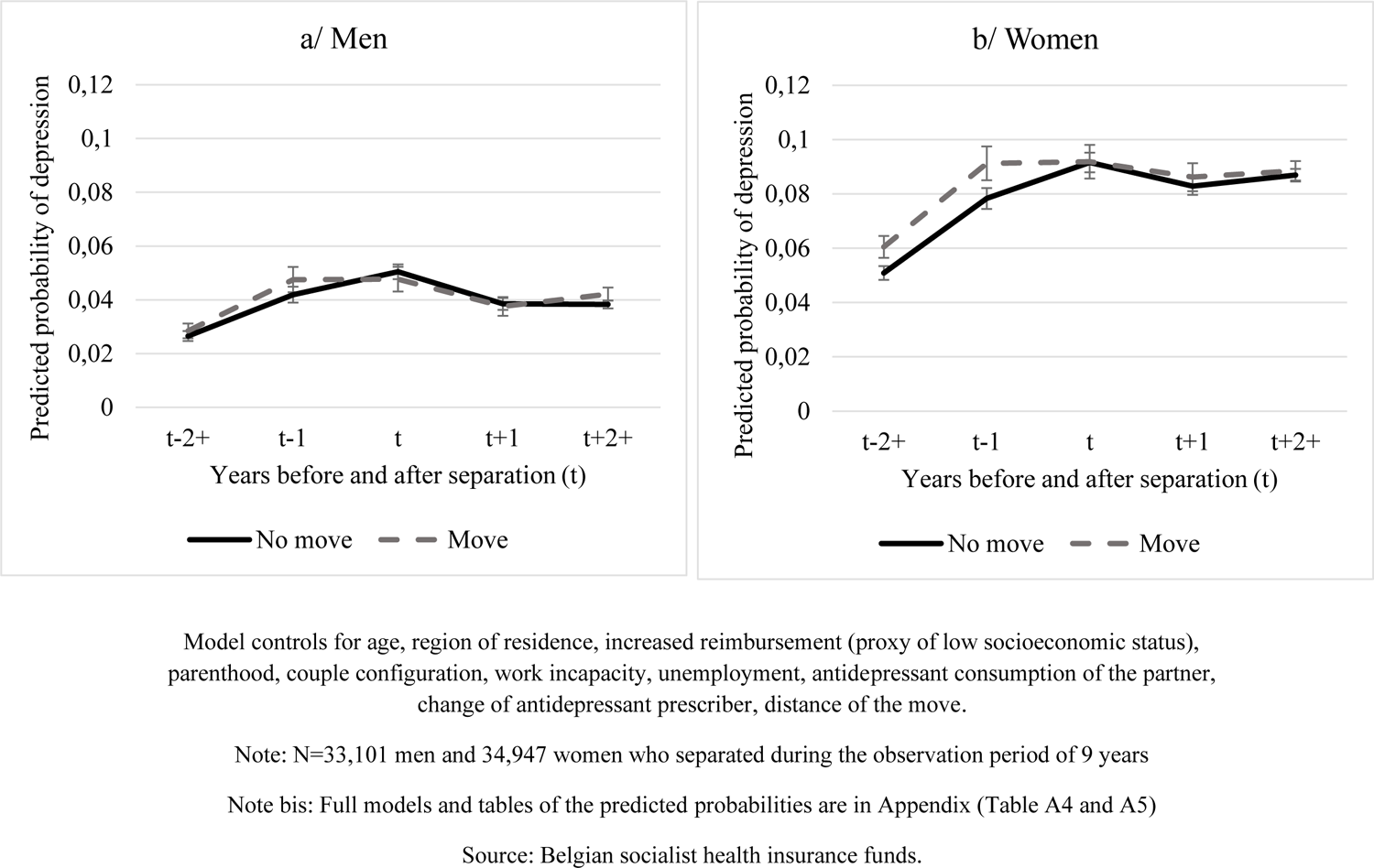
Predicted probability of antidepressants intake being 90DDD+ a year (based on random-effect logistic regression models) according to the mobility status of the individual at the time of the separation (t). The mobility status during the separation year (t) is attributed to the whole period (before and after the separation).

### Third hypothesis

Figure 4 shows the distinction between the individuals who will repartner after their separation and those who will remain single. Men do not present much difference in medication use patterns according to their moving status and repartnership trajectories after separation. Women who find a new partner quickly after the separation show a lower risk of antidepressants consumption, especially when they moved and repartnered in the same year. In the following year, women who are in a new relationship present lower medication use, no matter whether they moved or not. This confirms our third hypothesis: for women, moving in a context of separation and immediate repartnership is associated with lower intake than moving in a context of separation with no new partner. In the longer run, we can also confirm that repartnership is related to less antidepressants use for women, no matter the mobility pattern at the moment of the separation.

**Figure 4.**
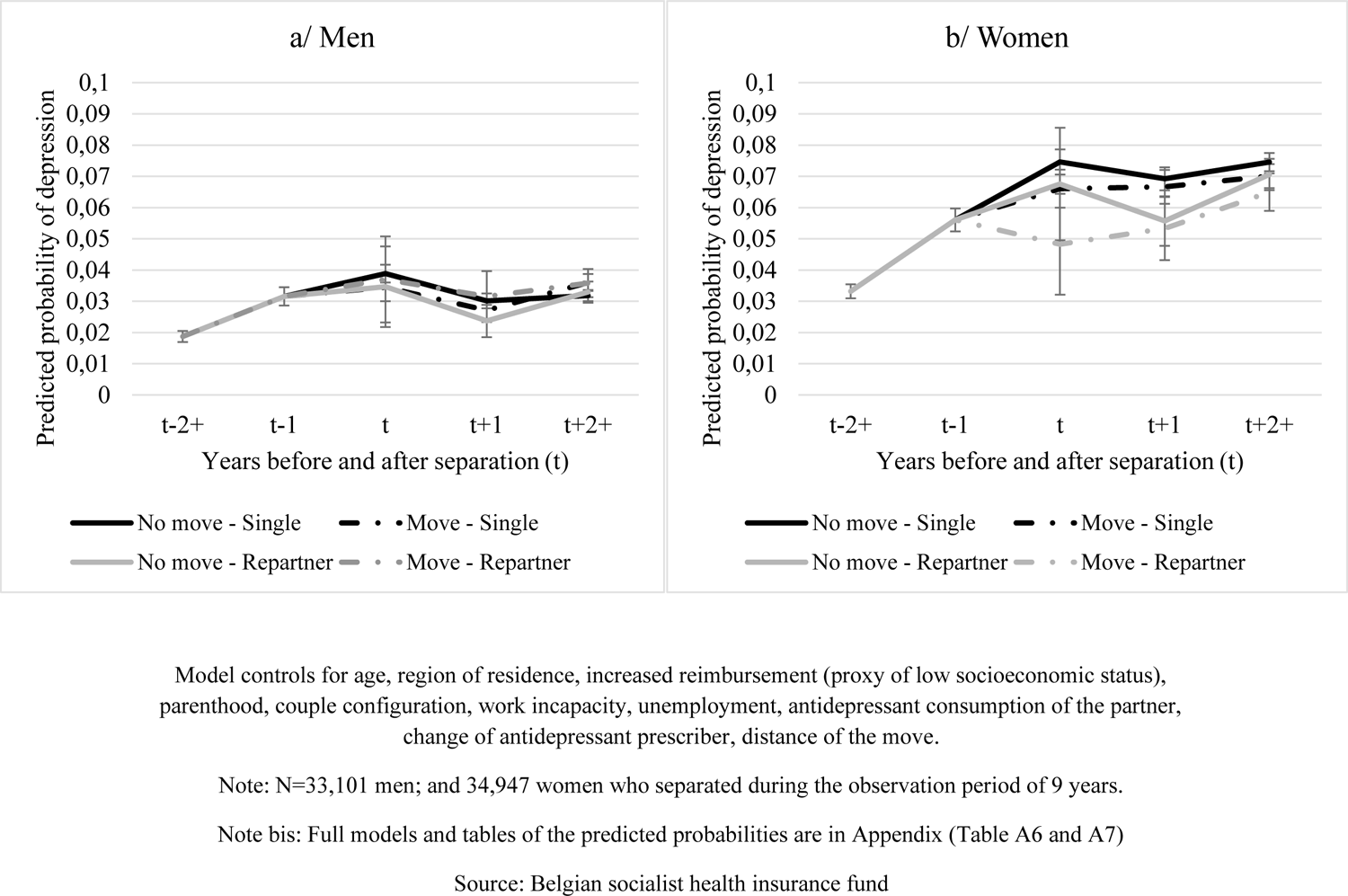
Predicted probabilities of antidepressants intake being 90DDD+ a year (based on logistic regression models) according to the mobility status of the individual at the moment of the separation (t) and the following year, and according to the repartnership status. The mobility status during the separation year (t) and the year after the separation is attributed from this separation. Source:

### Robustness checks

For hypotheses 1 and 2, for which results are not expected, some robustness checks were conducted to test to what extent the methodological choices could influence our results. First, we estimated the predicted margins of antidepressants intake in DDD – without any threshold – for individuals who moved or did not move in the year of separation according to the steps in the union dissolution process with linear regression models (Appendix - Figures A1 and A2). Second, depression remaining a rather rare event that concerns less than 10% of the population, we then conducted Poisson regression models – that are appropriate for rare events (Cai et al., 2010) – allowing us to calculate the incidence rate ratio of depression, defined by the threshold of 90 DDD and more per year (Appendix, Figures A3 and A4). In the two cases, previous observations for hypotheses 1 and 2 remain robust to these alterative specifications.

Third, we considered only couples in which both partners did not consume any antidepressants at the beginning of the observation, during the year 2008 (Figure 4). The objective is to remove all couples in which there is at least one individual who already consumed antidepressants and may suffer from chronic mental health issues, and thus to see whether the results are driven by a selectivity effect according to which a poorer mental health would lead to less stability both in terms of partnership and housing career. These checks support previous results, including when it comes to women’s higher predicted probability of antidepressants use during the whole observation period, in the case when they move out of the shared place during the year of the union dissolution (Figure A5 – b). However, women who did not consume any antidepressant at the beginning of the observation period present a lower medication use when they do not move during the year of separation (compared to moving in year t), especially at this exact moment. In addition, Figure A5 shows that the predicted probability for individuals to consume antidepressant at the beginning of the observation remains high two and more years after the separation for both men and women, no matter they leave or stay in the shared municipality of residence.

## CONCLUSIONS

This study aimed at investigating the role of residential mobility in the negative relation between union dissolutions and mental health. Drawing data from the socialist health insurance fund, we used a sample of couples who were affiliated to the SHIF, intact on January 1^st^ 2009 and separated in the 2009-2018 period. Applying random-effect logistic regression models, we estimated the probability of an antidepressant intake of at least 90 Defined Daily Doses per year, that is a treatment of at least 3 months over the year. Compared to the period of two and more years before the separation, the probability of medication sue starts to increase in the year prior to separation, is the highest during the separation year, and then decreases again (Table 2). Pre-separation levels are reached by men, whereas women present a higher probability of antidepressants use at two years and more after separation, compared to two and more years before the separation.

We observed that differences in antidepressants intake between persons who moved and who did not move (or changed municipalities) in the year of separation are very limited **(H1 not confirmed).** Residential mobility is surely a stressful and demanding time for the newly separated or divorced. First, it is – in most cases – a relatively forced move, made in emergency and with a pressure to find a convenient place as soon as possible after the decision is made to separate. Second, separation is accompanied with a decrease in resources at the level of the household. This can lead to a loss in housing conditions and neighbourhood satisfaction, which are related to negative mental health outcomes such as higher risk of depression (Evans et al., 2003; Singh et al., 2019). Third, if movers often face a lack of social support and less social integration than locals (Hendriks et al., 2016), this is particularly detrimental in a context of fragility that is the end of a relationship. Facing such a stressful and painful life event, the geographical distance can be an obstacle to the help of friends and family, and a marker of the distance with the children.

But on the other hand, mobility can also bring advantages and new opportunities (Mulder, 2018; Mulder & Van Ham, 2005; Trigg, 2009). First, it allows to physically disconnect from the ending relationship and start a new chapter in one’s life. Indeed, places are attached to feelings, and leaving can sometimes help overcome traumatic experiences (Trigg, 2009). Second, mobility is positively associated with a lot of outcomes, especially the professional life and the income (Mulder & Van Ham, 2005). Focusing on other achievements can be used as a coping mechanism for the newly separated. Third, the person who leaves the shared housing might also be the one who made the decision to leave the relationship. In such a situation, the mobility is not forced. To fully understand the implications of mobility in a context of union dissolution on one’s mental health and wellbeing, it is important to also understand the context of the separation, which is an information that is hard to find in administrative datasets. Future studies, especially qualitative studies, should focus on how separation-driven moves are experienced by individuals and how they can impact their psychological, material and social wellbeing.

We included a double approach, with both time-constant and time-varying mobility indicators in the separation year. This allowed to focus on possible anticipatory effects and to confirm a gender specificity in the relation between mobility in a context of union dissolution and antidepressants treatment **(H2 confirmed).** Women who moved at the time of separation presented a higher probability of medication intake during the whole observation period. This result points to a possible anticipation of the separation. Women tend to initiate the separation (Hewitt et al., 2006) more and persons who initiate the separation are likelier to move (Mulder & Wagner, 2010). In such a context, we can assume that women who will move out of the shared accommodation in the separation year are also more likely to have their mental state decrease before the separation and to have suffered from a poor-quality partnership before the separation. For instance, victims of psychologically and/or physically abusive relationships are likelier to suffer from depression, distress and to consume antidepressants (Ruiz-Pérez & Plazaola-Castaño, 2005). Women with fewer economic means might also be afraid of a separation as they know they will not be financially able to stay in the shared place, which can impact their happiness within the couple and after the separation as well as their power of decision in the separation (Aizer, 2010; Chesley, 2011; Malone et al., 2010). Another explanation could rely in a selectivity process: poor mental health can lead to a more instable life course (Afifi et al., 2006; Booth & Amato, 1991; Butterworth & Rodgers, 2008; Wade & Pevalin, 2004). Women with predisposition to poor mental health would show a higher risk to be mobile at the moment of the separation. This result also applies to women with did not consume antidepressant in 2008.

Findings point to an anticipation effect of union dissolution with respect to antidepressants consumption and to a gendered effect regarding the duration: for men, the impact of union dissolution is limited to the year of separation and the first year thereafter. For women, we identify a longer-term effect. This can be explained by the fact that the consequences of a separation among women tend to impact multiple parts of their lives, such as their risk of poverty and single motherhood (Fernquist & Cutright, 1998; Leopold, 2018). On the contrary, men especially face a (temporary) reduction of their social support system (Leopold, 2018). The specific operationalisation of mental health in this study could have impacted our results. The fast decrease of the depression risk among men might also be related to the fact that they seek less health care, especially when they are not partnered (Dykstra & Keizer, 2009; Payne et al., 2008). They also tend to present less healthy behaviour and less adherence to healthcare when they are unpartnered or separated than when they have a partner (Williams & Umberson, 2004). We expected men to present a higher intake of antidepressants when they change residences at the moment of the separation, as they may encounter more mental health struggles, and have less social support and related coping mechanisms to face this difficult life event. This is not the case. However, it is important to note that our way to measure depression – through a medication consumption – might not only reveal men’s mental health state, but also a lower tendency to seek for medical help when they are single (Dykstra & Keizer, 2009; Payne et al., 2008).

This gender specificity of the relation is confirmed by the analysis distinguishing individuals who entered a new relationship in the years following the separation from those who remained single. While men show little differences in probability of depression according to this variable, women who enter a new relationship during the separation year show a lower risk of depression, especially if they leave the previously shared place at this moment **(H4 confirmed for women**).

This might indicate that women in a new relationship can suffer less from the negative impact of separation especially when the move during the separation year happens in the context of a new partnership formation, compared to women who remain single. First, a certain share of women might have started their new relationship before or right after the first separation, which reduces the emotional suffering caused by the union dissolution (Crosier et al., 2007). Then, a new partnership might reduce the economic hardship related to the separation (Dziak et al., 2010; Leopold, 2018). For women, a separation is likelier to be associated with a loss of socioeconomic means and resources. A new partnership will decrease this risk by rising the resources and income at the household level. Finally, a selective process might also lead women with better mental health outcomes to start a new relationship faster (Wu & Hart, 2002). For men, this beneficial effect is less visible, which might be explained by the relation between men and healthcare. Men in a relationship can also tend to consume more antidepressants when they are surrounded, or in a relationship, where healthy habits are encouraged (Hughes & Waite, 2009; Wu & Hart, 2002).

### Limitations and methodological considerations

We could rely on a large-scale longitudinal dataset that result from the coupling of two high-quality datasets: the SHIF dataset to provide us medical information about the affiliated and the National Register for information about partnership transitions and residential mobilities. This dataset presents some limitations. First, our sample is limited to the members of the socialist health insurance fund, and more specifically to couples in which both partners are members of this insurance. This sample can be considered as biased compared to the general population. In Belgium, the choice of health insurance is based on family-related preferences, values and the self-defined position in society (D’hoore & Stordeur, 2004; Faniel & Gobin, 1992). Compared to the Belgian population, members of the socialist health insurance fund are on average more deprived – as shown by a higher number of persons with increased reimbursement (see Appendix – Table A1). The couples in our sample are also more often unmarried and childless, which is associated with a higher partnership instability (Musick & Michelmore, 2018). Finally, future studies could profit from increasing access to detailed administrative data, including e.g. more detailed information about dates and events which would allow to better draw causal conclusions on the effect of life event on mental health. So far, in Belgium, the merging of medical data and register data is very rare and framed for data privacy reasons, and we cannot estimate to what extent this selectivity in our sample biases the results.

The choice of antidepressants as a proxy for mental health is usual in the specialised literature. This is a limited proxy for mental health itself, as it hides possible undiagnosed mental health struggles and depression. However, in a context such as a separation, sadness and grief are two normal mechanisms. Using a medical and quantifiable tool, such as antidepressants intake remove the risk of subjective assessments. The results of a wellbeing scale or a list of psychological symptoms would highly depend on the context of the study – the time since the separation, the conflicts between the ex-partners at the moment of the interview – while yearly antidepressants intake summarises the mental health challenges happening during this life period and gives an objective measurement, based on a doctor’s diagnosis.

Another element worth mentioning is the absence of distinction we made between marital and non-marital unions. The determinants of marriage, compared to cohabiting relationships, are related to socioeconomic characteristics. Women with low educational attainment have less stable partnership and family trajectories; they are likelier to enter cohabiting relationship early in life, tend less to transition to marriage, and when they do, they have a higher divorce risk (Lundberg et al., 2016). Because we cannot control for education or occupation, and we cannot explain the possible differences between marital and nonmarital unions, we decided to consider all unions the same way.

Our models accounted for individuals’ characteristics such as age, sex, household composition and region of residence. They also included unemployment, financial precariousness and work incapacity, which are determinants of individual’s wellbeing and mental health (Conejero et al., 2018; Lersch & Vidal, 2014). Nevertheless, other elements are relevant in the relation between separation, mobility and mental health and were not considered in our models. We control for increased reimbursement that is offered to low-income individuals, that was used in previous studies as a relevant proxy for socioeconomic status (Van den Bosch et al., 2013), but we have little knowledge on the deprivation level or variation in earnings among the population. A limitation of this study is that standard socioeconomic indicators, such as educational attainment or income, are lacking in our database. Controlling for this information would possibly disclose a more negative impact of union dissolution on men’s mental health (Feijten, 2005; Wyder et al., 2009). However, the use of random-effect models helped to control for individuals’ specificity and unobserved characteristics, such as biological determinants or cultural background.

### Contribution

Despite some limitations, we consider this insurance-based panel data as a real asset in the observation of mental health changes during life course events, such as separation. Our article presents two important contributions to literature. First, it highlights the complex relation between separation-driven mobility and mental health and the existence of both advantages and disadvantages, of staying in the previous place rather than moving out. It calls for a more in-depth analysis of the contextual elements of the relationship dissolution that could be associated with the moving decision, such as who initiates the separation, the level of conflicts at the moment of the separation and over the children custody, the personal resources and income. Second, it contributes to a better understanding of the gender-specific consequences of separations and separation-driven residential mobility on mental health. Our findings suggest that women who leave the shared place face a lower mental health before the separation. This highlights the detrimental impact of low-quality relationships on women’s mental health and their anticipation of the separation and relocation, especially in a context where women suffer the most from long-term economic losses after a divorce or a separation.

These conclusions encourage direct actions. We recommend public policies support individuals at two times. First, at the moment of the separation. Low-quality relationships are detrimental for mental health – we observed it for women especially – and policies should help individuals in finding and affording a new residence, that is adapted to their needs and their family’s needs. Services of relocation for women leaving a difficult (or even abusive) relationship should be encouraged. Second, in the longer run after the separation. For women especially, a divorce or separation is a time of social and economic fragility. We can see that repartnering is often considered as a protective factor against poverty and income decrease for women, and policies should offer an alternative to this by supporting single mothers and newly separated women quickly after the separation, but also in the longer term. Finally, this article recalls that men tend to consume less antidepressants than women, in a society where men are likelier to end due to suicide, especially after they separate or divorce (Bruce & Kim, 1992; Payne et al., 2008). Policies and societal debate about men’ mental health should target a change of mentalities and behaviours, and help men seek medical help open up about their inner struggles.

## Data Availability

All data produced in the present work cannot be shared. Their protection is ensured by the Socialist Health Insurance Funds.

## APPENDIX

**Table A-1.** Random-effect logit regression on the risk of consuming at least 90DDD of antidepressants over a year for men and women, expressed in Odds Ratio.

|  | Men |  | Women |  |
| --- | --- | --- | --- | --- |
|  | Model 1 | Model 2 | Model 1 | Model 2 |
| N | 49,930 | 49,930 | 52,786 | 52,786 |
| Separation period |  |  |  |  |
| 2 years and more before separation | 1.00 | 1.00 | 1.00 | 1.00 |
| 1 year before separation (t-1) | 1.63<br>[1.52-1.75] | 1.63<br>[1.52-1.75] | 1.58<br>[1.51-1.68] | 1.58<br>[1.51-1.68] |
| Year of separation (t) | 1.80<br>[1.67-1.95] | 1.80<br>[1.65-1.95] | 1.84<br>[1.73-1.95] | 1.82<br>[1.71-1.93] |
| 1 year after separation (t+1) | 1.13<br>[1.04-1.22] | 1.13<br>[1.03-1.22] | 1.46<br>[1.38-1.55] | 1.45<br>[1.37-1.54] |
| 2 years and more after separation | 0.99<br>[0.92-1.06] | 0.99<br>[0.93-1.76] | 1.22<br>[1.16-1.30] | 1.21<br>[1.14-1.29] |
| Residential move at t |  | 1.01<br>[0.94-1.08] |  | 1.02<br>[0.98-1.08] |
| Age | 1.06<br>[1.05-1.06] | 1.06<br>[1.05-1.06] | 1.09<br>[1.09-1.10] | 1.09<br>[1.09-1.10] |
| Increased reimbursement | 1.43<br>[1.34-1.52] | 1.43<br>[1.34-1.52] | 1.30<br>[1.23-1.35] | 1.30<br>[1.23-1.35] |
| Parent of resident children | 0.96<br>[0.90-1.02] | 0.96<br>[0.90-1.02] | 1.04<br>[1.00-1.08] | 1.04<br>[1.00-1.08] |
| Married or in non-marital relationship | 0.89<br>[0.86-0.92] | 0.89<br>[0.86-0.92] | 0.92<br>[0.90-0.95] | 0.92<br>[0.90-0.95] |
| Region (ref. Flanders) |  |  |  |  |
| Brussels | 0.86<br>[0.74-0.98] | 0.86<br>[0.74-0.98] | 0.68<br>[0.61-0.77] | 0.68<br>[0.61-0.77] |
| Wallonia | 1.72<br>[1.58-1.86] | 1.72<br>[1.58-1.86] | 2.10<br>[1.97-2.25] | 2.10<br>[1.97-2.25] |
| Incapacity | 3.25<br>[2.97-3.53] | 3.25<br>[2.97-3.53] | 2.86<br>[2.69-3.03] | 2.86<br>[2.69-3.03] |
| Unemployment | 1.03<br>[1.90-1.15] | 1.03<br>[1.90-1.15] | 1.01<br>[0.90-1.14] | 1.01<br>[0.90-1.14] |
| Consumption of the partner | 1.01<br>[1.01-1.01] | 1.01<br>[1.01-1.01] | 1.00<br>[1.00-1.01] | 1.00<br>[1.00-1.01] |
| Change of prescriber |  |  |  |  |
|  | 4.81<br>[4.62-5.00] | 4.81<br>[4.62-5.00] | 2.80<br>[2.72-2.89] | 2.80<br>[2.72-2.89] |
| Intercept | -8.59 | -8.59 | -8.95 | -8.95 |

Note: N=33,101 (297,909 person-years); and 34,947 women (314,523 person-years), who separated during the observation period of 9 years.
Control for age, region of residence, increased reimbursement (proxy of low socioeconomic status), parent of resident children (vs. childless or parent of non-resident children), in a married or non-married relation (vs. living without a partner), work incapacity (more than 150 days of sick leave a year), unemployment (more than 150 days of unemployment a year), antidepressant consumption of the partner, change of antidepressant prescriber.
Source: Belgian socialist health insurance fund.
Note: Depression is measured by the consumption of at least 90DDD of antidepressants

### Hypothesis 1

**Tableau A-2.** Random-effect logit regression on the risk of consuming at least 90DDD of antidepressants over a year for men and women, expressed in Odds Ratio.

|  | Men |  |  | Women |  |  |
| --- | --- | --- | --- | --- | --- | --- |
|  | OR | CI 95% |  | OR | CI95% |  |
| Timing of the separation x Mobility at the moment of the separation<br>(ref. 2 years+ before separation x No move) |  |  |  |  |  |  |
| 1 year before separation x No move | 2.05 | 1.82 | 2.30 | 2.14 | 1.95 | 2.33 |
| Year of separation x No move | 2.77 | 2.43 | 3.16 | 2.86 | 2.59 | 3.17 |
| Year after separation x No move | 1.78 | 1.56 | 2.03 | 2.37 | 2.15 | 2.62 |
| 2 years+ after separation x No move | 1.77 | 1.57 | 1.98 | 2.59 | 2.37 | 2.83 |
| 2 years+ before separation x Move | 1.11 | 0.94 | 1.31 | 1.35 | 1.18 | 1.53 |
| 1 year before separation x Move | 2.51 | 2.10 | 3.01 | 2.84 | 2.46 | 3.28 |
| Year of separation x Move | 2.52 | 2.07 | 3.08 | 2.88 | 2.46 | 3.37 |
| Year after separation x Move | 1.71 | 1.42 | 2.06 | 2.55 | 2.21 | 2.94 |
| 2 years+ after separation x Move | 2.06 | 1.78 | 2.39 | 2.68 | 2.39 | 3.01 |
| Age | 1.03 | 1.03 | 1.04 | 1.05 | 1.05 | 1.06 |
| Preferential fees (ref. No) | 1.42 | 1.30 | 1.55 | 1.26 | 1.19 | 1.33 |
| Have children | 0.99 | 0.92 | 1.07 | 1.12 | 1.06 | 1.17 |
| Couple status (ref. Unpartnered) |  |  |  |  |  |  |
| Marital union | 0.88 | 0.79 | 0.98 | 0.84 | 0.77 | 0.90 |
| Non-marital union | 0.80 | 0.73 | 0.88 | 0.85 | 0.79 | 0.91 |
| Region of residence (ref. Flanders) |  |  |  |  |  |  |
| Brussels | 0.73 | 0.63 | 0.86 | 0.75 | 0.66 | 0.86 |
| Wallonia | 1.46 | 1.34 | 1.60 | 1.60 | 1.49 | 1.72 |
| Temporary work incapacity | 3.23 | 2.91 | 3.59 | 2.96 | 2.75 | 3.19 |
| Long-term work incapacity | 6.82 | 6.20 | 7.49 | 5.61 | 5.24 | 6.01 |
| Temporary unemployment | 0.89 | 0.79 | 1.01 | 0.97 | 0.88 | 1.05 |
| Long-term unemployment | 0.99 | 0.86 | 1.14 | 0.95 | 0.86 | 1.05 |
| Partner's antidepressants consumption | 1.00 | 1.00 | 1.00 | 1.00 | 1.00 | 1.00 |
| Change of prescriber | 9.62 | 9.12 | 10.15 | 5.05 | 4.86 | 5.24 |
| Distance of the move (ref. No move) |  |  |  |  |  |  |
| Under 16 km | 1.07 | 0.96 | 1.18 | 1.07 | 0.99 | 1.15 |
| 16km and more | 1.00 | 0.88 | 1.14 | 0.95 | 0.85 | 1.06 |
| Constant | 0.00 | 0.00 | 0.00 | 0.00 | 0.00 | 0.00 |
| Panel-level variance component | 1.77 | 1.72 | 1.82 | 1.89 | 1.85 | 1.92 |
| Standard deviation | 2.43 | 2.37 | 2.49 | 2.57 | 2.52 | 2.61 |
| Proportion of the total variance contributed by the panel-level variance component | 0.64 | 0.63 | 0.65 | 0.67 | 0.66 | 0.67 |

**Tableau A-3.** Predicted probabilities (PP) of consuming at least 90DDD of antidepressants over a year for men and women, according to whether men and women moved during the separation year or not. Moving status is attributed to the whole period (before and after the separation/mobility).

| Men |  |  |  |  |  |  |
| --- | --- | --- | --- | --- | --- | --- |
|  | No move |  |  | Move |  |  |
|  | PP | IC95% |  | PP | IC95% |  |
| 2 years+ before separation | 0.027 | 0.025 | 0.028 | 0.028 | 0.026 | 0.031 |
| Year before separation | 0.042 | 0.039 | 0.045 | 0.048 | 0.043 | 0.052 |
| Year of separation | 0.050 | 0.048 | 0.053 | 0.048 | 0.043 | 0.052 |
| Year after separation | 0.038 | 0.036 | 0.041 | 0.038 | 0.034 | 0.041 |
| 2 years+ after separation | 0.038 | 0.037 | 0.040 | 0.042 | 0.040 | 0.045 |
| Women |  |  |  |  |  |  |
|  | No move |  |  | Move |  |  |
|  | PP | IC95% |  | PP | IC95% |  |
| 2 years+ before separation | 0.051 | 0.048 | 0.053 | 0.060 | 0.056 | 0.065 |
| Year before separation | 0.078 | 0.074 | 0.082 | 0.091 | 0.085 | 0.098 |
| Year of separation | 0.092 | 0.088 | 0.095 | 0.092 | 0.086 | 0.098 |
| Year after separation | 0.083 | 0.080 | 0.086 | 0.086 | 0.081 | 0.091 |
| 2 years+ after separation | 0.087 | 0.085 | 0.089 | 0.088 | 0.085 | 0.092 |

### Hypothesis 2

**Tableau A-4.** Random-effect logit regression on the risk of consuming at least 90DDD of antidepressants over a year for men and women, expressed in Odds Ratio.

|  | OR | Men<br>*CI 95%* |  | OR | Women<br>*CI95%* |  |
| --- | --- | --- | --- | --- | --- | --- |
| Timing of the separation x Mobility at the moment of the separation<br>(ref. 2 years+ before separation x No move) |  |  |  |  |  |  |
| 1 year before separation x No move | 2.11 | *1.91* | *2.33* | 2.15 | *1.99* | *2.31* |
| Year of separation x No move | 2.76 | *2.46* | *3.09* | 2.90 | *2.66* | *3.17* |
| Year after separation x No move | 1.77 | *1.58* | *1.99* | 2.39 | *2.19* | *2.61* |
| 2 years+ after separation x No move | 1.75 | *1.58* | *1.93* | 2.59 | *2.40* | *2.79* |
| Year of separation x Move | 2.38 | *2.00* | *2.83* | 2.54 | *2.22* | *2.91* |
| Year after separation x Move | 1.62 | *1.38* | *1.90* | 2.27 | *2.02* | *2.55* |
| 2 years+ after separation x Move | 1.96 | *1.74* | *2.22* | 2.42 | *2.21* | *2.65* |
| Age | 1.03 | *1.03* | *1.04* | 1.05 | *1.05* | *1.06* |
| Preferential fees (ref. No) | 1.42 | *1.30* | *1.55* | 1.26 | *1.19* | *1.34* |
| Have children | 0.99 | *0.92* | *1.07* | 1.10 | *1.05* | *1.15* |
| Couple | 0.90 | *0.85* | *0.94* | 0.93 | *0.90* | *0.96* |
| Region of residence (ref. Flanders) |  |  |  |  |  |  |
| Brussels | 0.74 | *0.63* | *0.86* | 0.75 | *0.66* | *0.85* |
| Wallonia | 1.46 | *1.34* | *1.60* | 1.60 | *1.49* | *1.73* |
| Temporary work incapacity | 3.23 | *2.91* | *3.59* | 2.96 | *2.74* | *3.19* |
| Long-term work incapacity | 6.82 | *6.20* | *7.49* | 5.60 | *5.23* | *6.00* |
| Temporary unemployment | 0.89 | *0.79* | *1.01* | 0.97 | *0.89* | *1.06* |
| Long-term unemployment | 0.99 | *0.86* | *1.14* | 0.95 | *0.86* | *1.05* |
| Partner's antidepressants consumption | 1.00 | *1.00* | *1.00* | 1.00 | *1.00* | *1.00* |
| Change of prescriber | 9.62 | *9.12* | *10.15* | 5.05 | *4.86* | *5.24* |
| Distance of the move (ref. No move) |  |  |  |  |  |  |
| Under 16 km | 1.07 | *0.97* | *1.18* | 1.07 | *0.99* | *1.15* |
| 16km and more | 1.00 | *0.88* | *1.14* | 0.95 | *0.85* | *1.06* |
| Constant | 0.00 | *0.00* | *0.00* | 0.00 | *0.00* | *0.00* |
| Panel-level variance component | 1.77 | *1.72* | *1.82* | 1.89 | *1.85* | *1.92* |
| Standard deviation | 2.43 | *2.37* | *2.49* | 2.57 | *2.52* | *2.61* |
| Proportion of the total variance contributed<br>by the panel-level variance component | 0.64 | *0.63* | *0.65* | 0.67 | *0.66* | *0.67* |

**Tableau A-5.** Predicted probabilities (PP) of consuming at least 90DDD of antidepressants over a year for men and women, according to whether men and women moved during the separation year or not. Moving status is attributed from the year of separation.

| Men |  |
| --- | --- |
| No move | Move |

|  | PP | IC95% |  | PP | IC95% |  |
| --- | --- | --- | --- | --- | --- | --- |
| 2 years+ before separation | 0.027 | 0.025 | 0.028 | 0.027 | 0.025 | 0.028 |
| Year before separation | 0.043 | 0.041 | 0.046 | 0.043 | 0.041 | 0.046 |
| Year of separation | 0.051 | 0.048 | 0.054 | 0.047 | 0.042 | 0.051 |
| Year after separation | 0.039 | 0.037 | 0.041 | 0.037 | 0.033 | 0.040 |
| 2 years+ after separation | 0.039 | 0.037 | 0.040 | 0.041 | 0.039 | 0.044 |

| Women |  |  |  |  |  |  |
| --- | --- | --- | --- | --- | --- | --- |
|  | No move |  |  | Move |  |  |
|  | PP | IC95% |  | PP | IC95% |  |
| 2 years+ before separation | 0.052 | 0.050 | 0.054 | 0.052 | 0.050 | 0.054 |
| Year before separation | 0.080 | 0.077 | 0.084 | 0.080 | 0.077 | 0.084 |
| Year of separation | 0.094 | 0.091 | 0.098 | 0.088 | 0.082 | 0.094 |
| Year after separation | 0.085 | 0.082 | 0.088 | 0.083 | 0.078 | 0.088 |
| 2 years+ after separation | 0.089 | 0.087 | 0.091 | 0.086 | 0.083 | 0.089 |

### Hypothesis 3

**Tableau A-6.** Random-effect logit regression on the risk of consuming at least 90DDD of antidepressants over a year for men and women, expressed in Odds Ratio.

|  | Men |  |  | Women |  |  |
| --- | --- | --- | --- | --- | --- | --- |
|  | OR | CI 95% |  | OR | CI95% |  |
| Timing of the separation x Mobility at the moment of the separation<br>(ref. 2 years+ before separation) |  |  |  |  |  |  |
| 1 year before separation | 2.16 | 1.90 | 2.47 | 2.31 | 2.09 | 2.56 |
| Year of separation x No move x Single | 2.99 | 2.47 | 3.61 | 3.79 | 3.28 | 4.38 |
| Year after separation x No move x Single | 2.02 | 1.66 | 2.45 | 3.32 | 2.87 | 3.84 |
| 2 years+ after separation x No move x Single | 2.19 | 1.83 | 2.62 | 3.78 | 3.30 | 4.33 |
| Year of separation x Move x Single | 2.47 | 1.92 | 3.19 | 3.06 | 2.52 | 3.72 |
| Year after separation x Move x Single | 1.72 | 1.35 | 2.19 | 3.11 | 2.60 | 3.72 |
| 2 years+ after separation x Move x Single | 2.65 | 2.17 | 3.23 | 3.39 | 2.92 | 3.95 |
| Year of separation x No move x Repartner | 2.50 | 1.42 | 4.40 | 3.18 | 2.02 | 5.02 |
| Year after separation x No move x Repartner | 1.41 | 1.03 | 1.92 | 2.29 | 1.82 | 2.88 |
| 2 years+ after separation x No move x Repartner | 2.31 | 1.99 | 2.68 | 3.44 | 3.06 | 3.86 |
| Year of separation x Move x Repartner | 2.76 | 1.56 | 4.90 | 1.81 | 1.05 | 3.10 |
| Year after separation x Move x Repartner | 2.16 | 1.47 | 3.18 | 2.13 | 1.56 | 2.89 |
| 2 years+ after separation x Move x Repartner | 2.63 | 2.18 | 3.17 | 2.97 | 2.56 | 3.46 |
| Age | 1.03 | 1.02 | 1.03 | 1.05 | 1.04 | 1.05 |
| Preferential fees (ref. No) | 1.39 | 1.26 | 1.54 | 1.23 | 1.15 | 1.31 |
| Have children | 0.98 | 0.90 | 1.07 | 1.11 | 1.05 | 1.18 |
| Couple | 0.90 | 0.81 | 0.99 | 0.99 | 0.91 | 1.07 |
| Region of residence (ref. Flanders) |  |  |  |  |  |  |
| Brussels | 0.71 | 0.60 | 0.83 | 0.77 | 0.67 | 0.88 |
| Wallonia | 1.44 | 1.31 | 1.58 | 1.54 | 1.42 | 1.68 |
| Temporary work incapacity | 3.01 | 2.67 | 3.39 | 2.98 | 2.73 | 3.26 |
| Long-term work incapacity | 6.63 | 5.96 | 7.39 | 5.63 | 5.19 | 6.10 |
| Temporary unemployment | 0.85 | 0.74 | 0.98 | 0.99 | 0.89 | 1.10 |
| Long-term unemployment | 0.97 | 0.82 | 1.14 | 0.89 | 0.79 | 1.00 |
| Partner's antidepressants consumption | 1.00 | 1.00 | 1.00 | 1.00 | 1.00 | 1.00 |
| Change of prescriber | 12.99 | 12.21 | 13.83 | 6.70 | 6.40 | 7.01 |
| Distance of the move (ref. No move) |  |  |  |  |  |  |
| Under 16 km | 1.04 | 0.93 | 1.17 | 1.08 | 0.99 | 1.18 |
| 16km and more | 1.06 | 0.91 | 1.23 | 0.96 | 0.85 | 1.09 |
| Constant | 0.00 | 0.00 | 0.00 | 0.00 | 0.00 | 0.00 |
| Panel-level variance component | 1.63 | 1.58 | 1.69 | 1.77 | 1.73 | 1.81 |
| Standard deviation | 2.26 | 2.20 | 2.32 | 2.42 | 2.37 | 2.47 |
| Proportion of the total variance contributed by the panel-level variance component | 0.61 | 0.59 | 0.62 | 0.64 | 0.63 | 0.65 |

**Tableau A-7.**
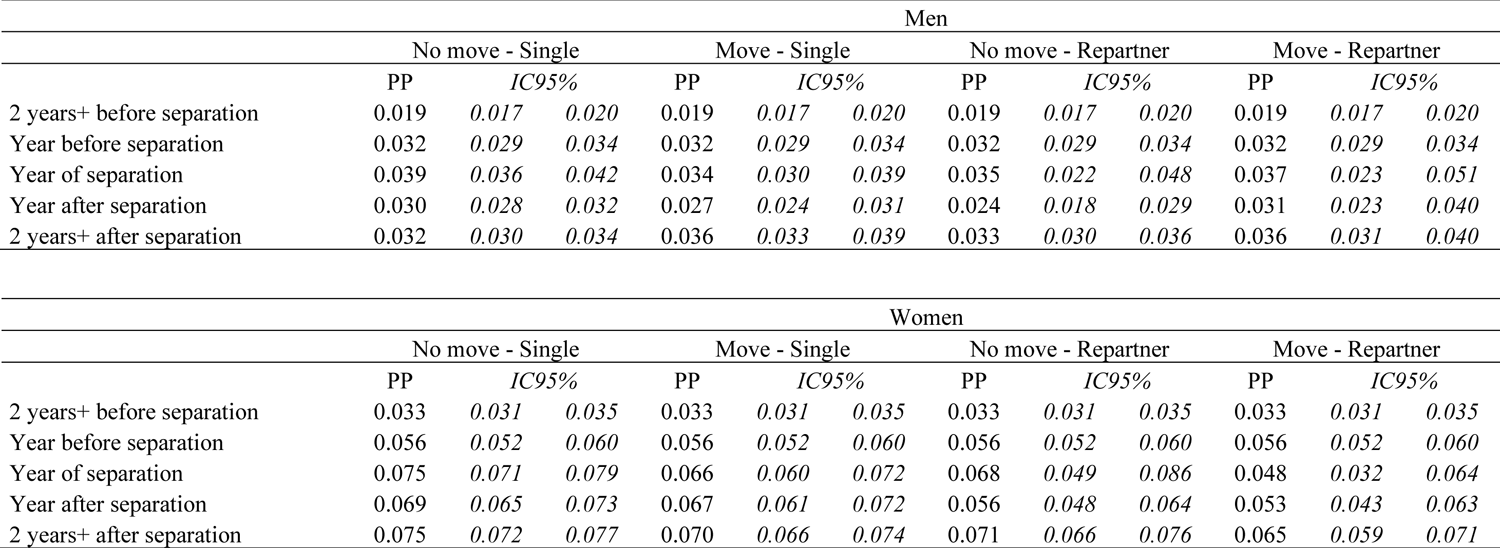
Predicted probabilities (PP) of consuming at least 90DDD of antidepressants over a year for men and women, according to whether men and women moved during the separation year or not, and their partnership status (single or repartnered).

### Robustness checks – Hypothesis 1

#### OLS models

##### Table margins

**Figure A1.**
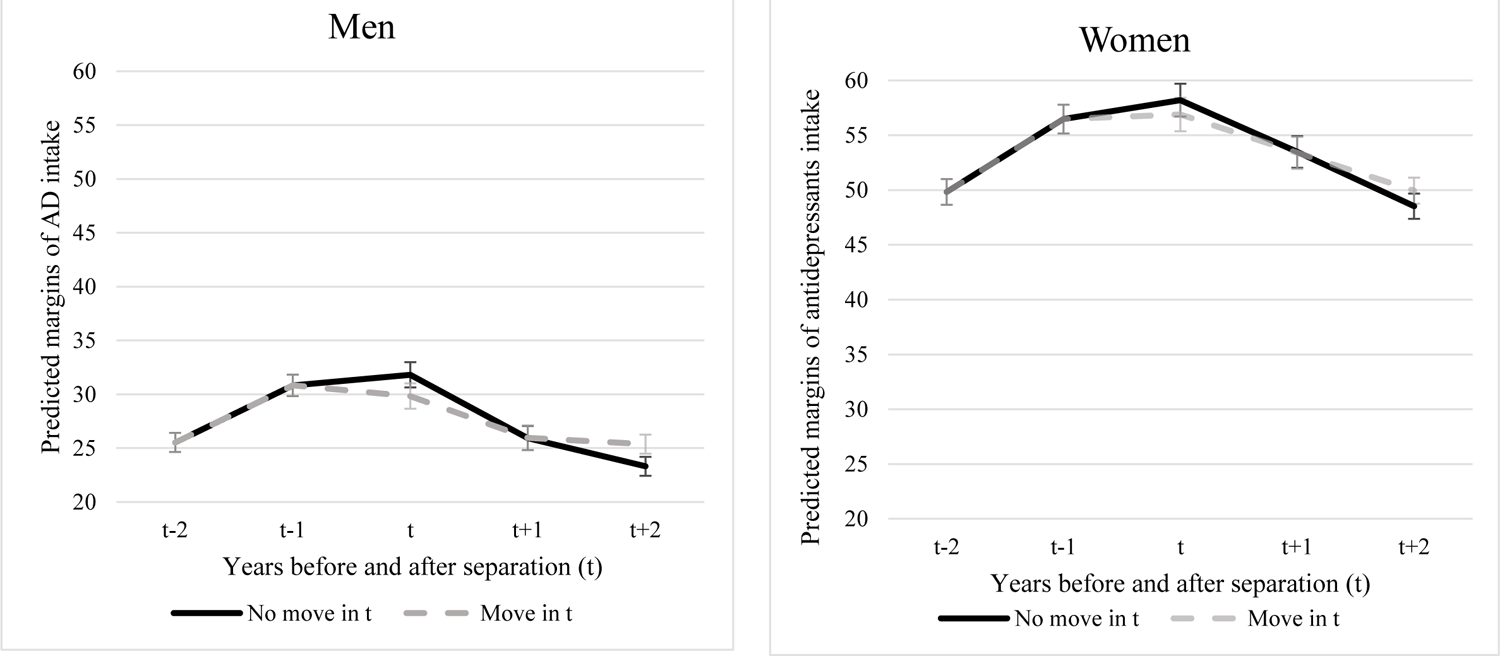
Predicted margins of antidepressant intake (based on OLS regression models) according to the mobility status of the individual at the moment of the separation (t). The mobility status during the year of separation (t) is attributed from this separation year (not before). Source: Belgian socialist health insurance fund.

**Figure A2.**
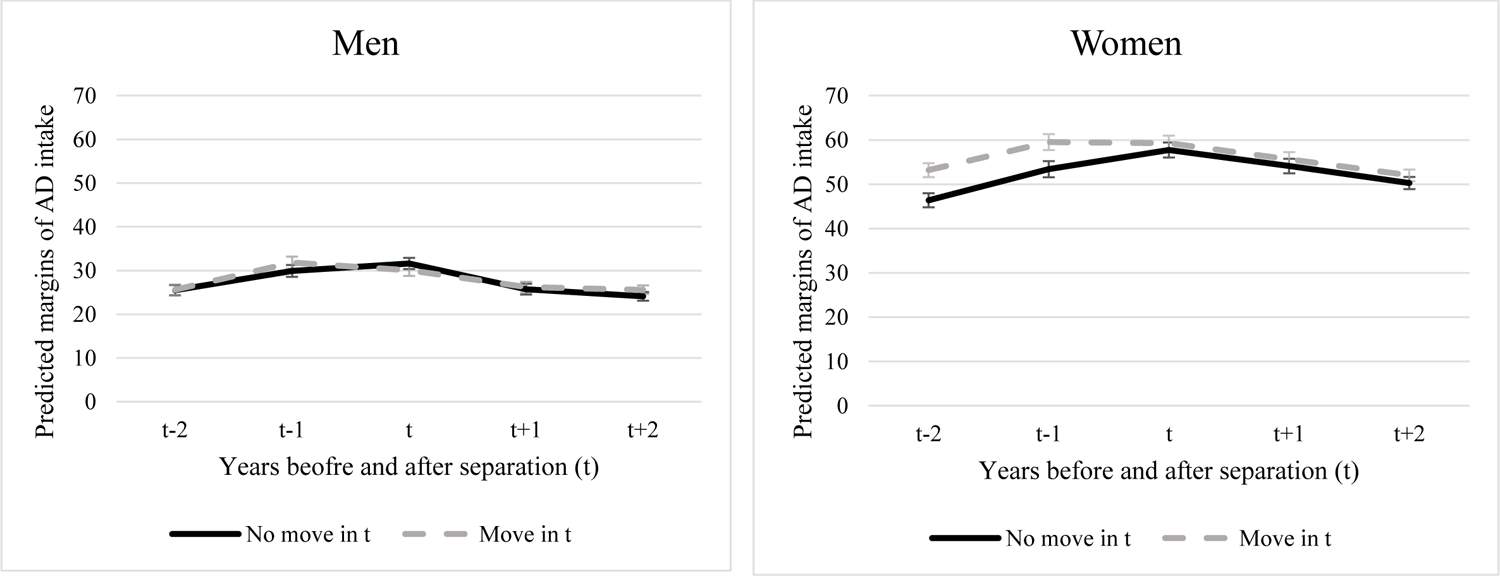
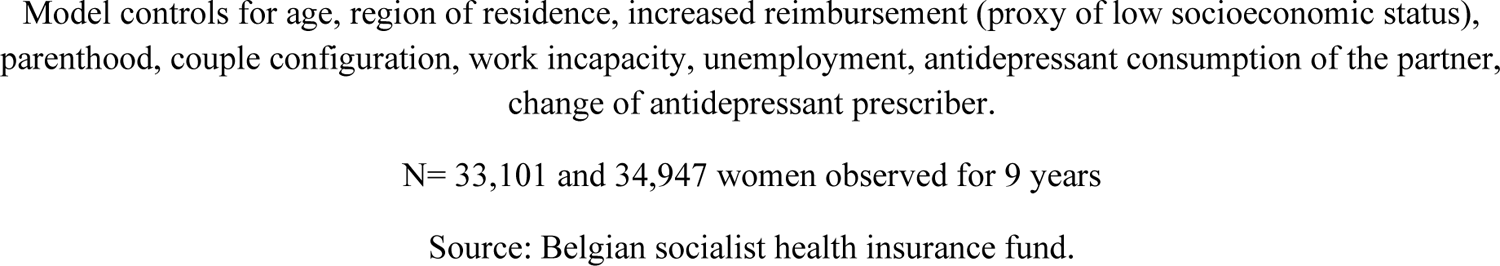
Predicted margins of antidepressant intake (based on OLS regression models) according to the mobility status of the individual at the moment of the separation (t). The mobility status during the year of separation (t) is attributed to the whole period (before and after the mobility). Source: Belgian socialist health insurance fund.

### Poisson models

**Figure A3.**
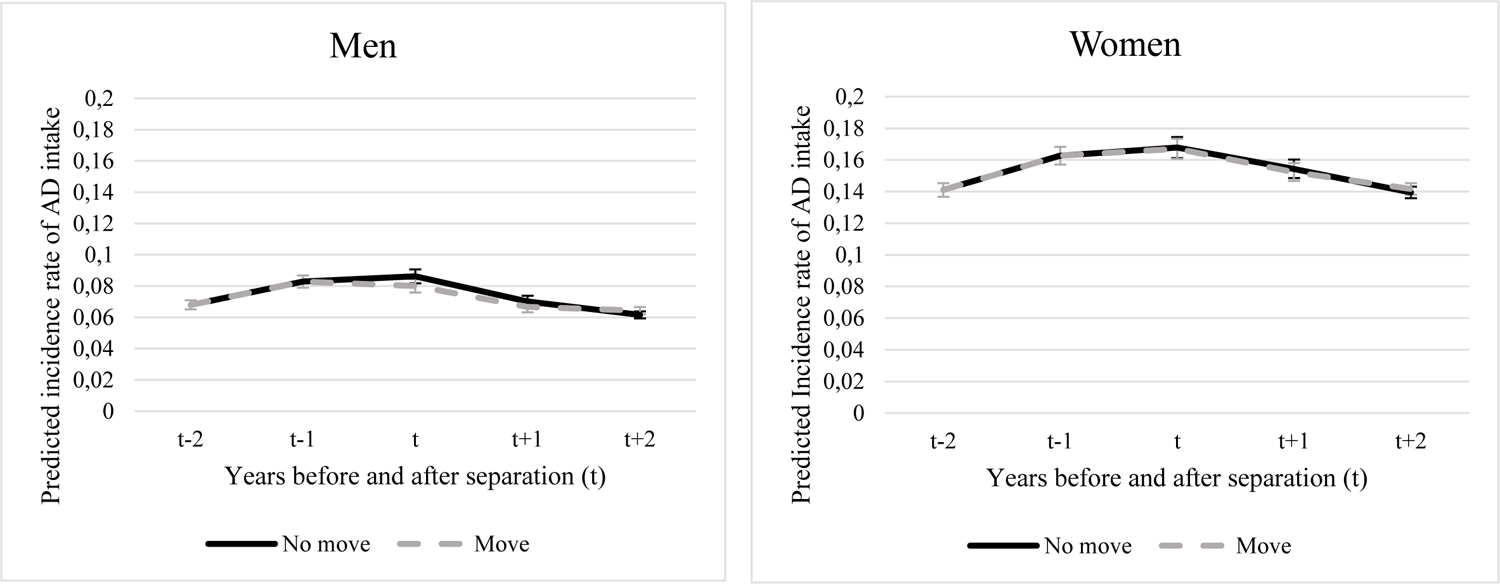
Predicted incidence rates of depression (based on Poisson regression models) according to the mobility status of the individual at the moment of the separation (t). The mobility status during the year of separation (t) is attributed from this separation year (not before). Source: Belgian socialist health insurance fund.

**Figure A4.**
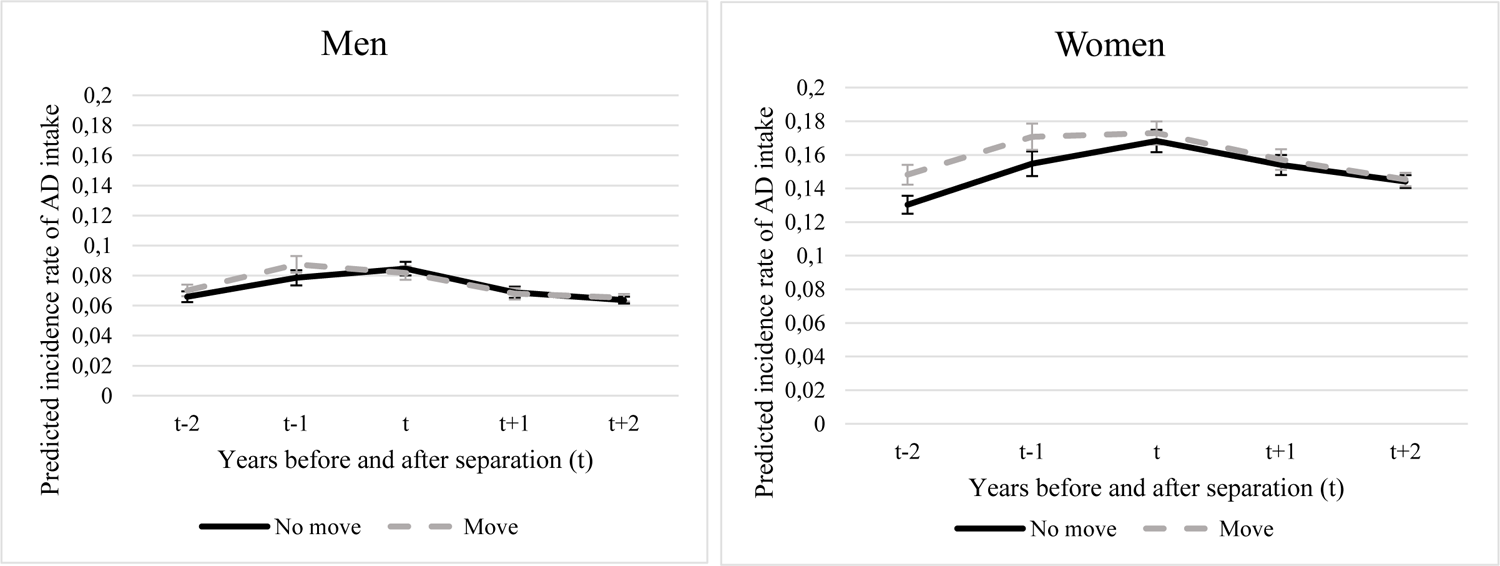
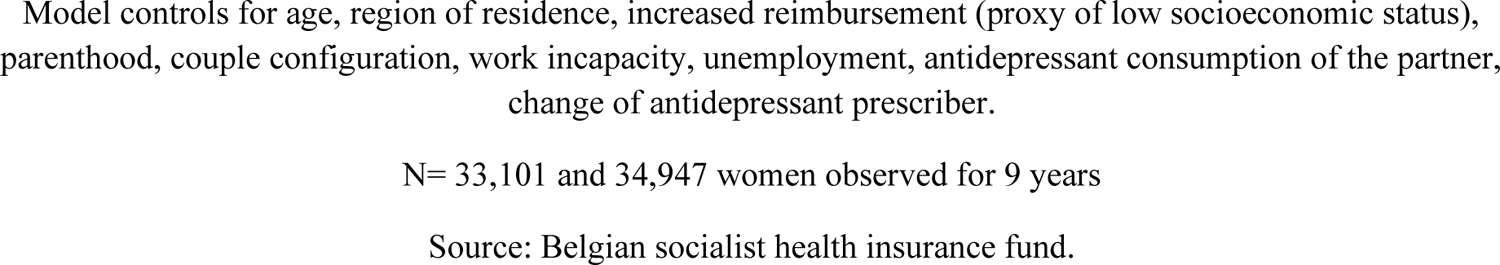
Predicted incidence rates of depression (based on Poisson regression models) according to the mobility status of the individual at the moment of the separation (t). The mobility status during the year of separation (t) is attributed during the whole period (before and after the mobility). Source: Belgian socialist health insurance fund.

### No consumption at the beginning of observation (in 2008)

**Figure A5.**
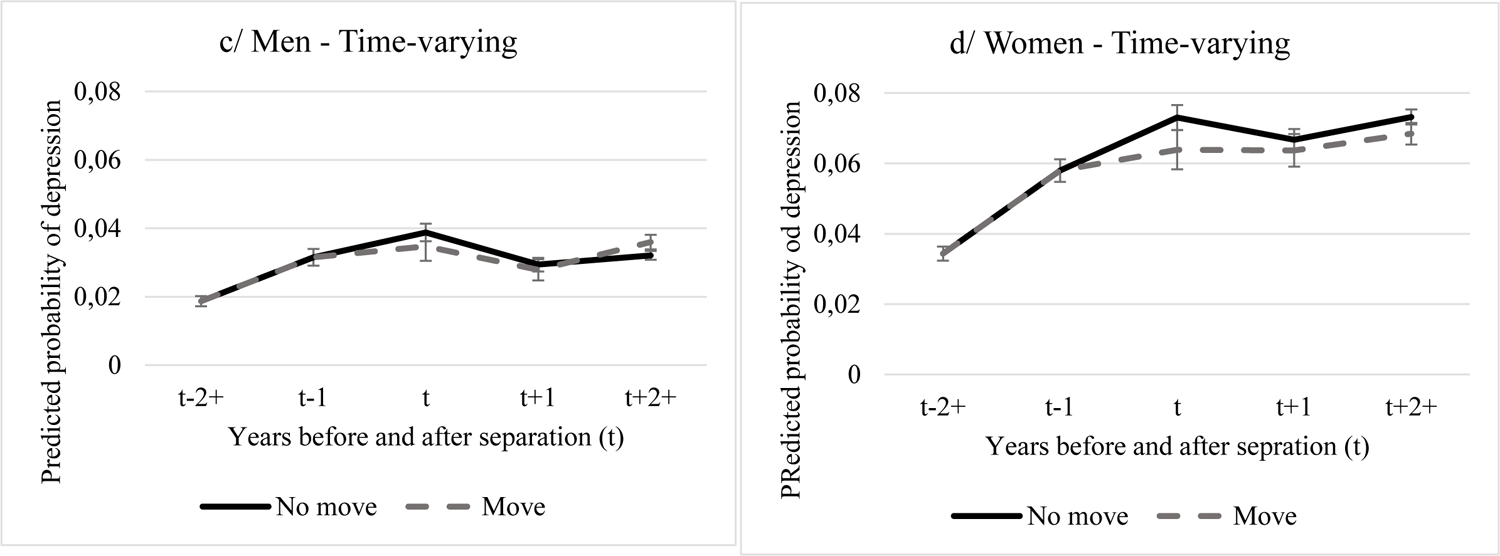
Predicted probabilities of a yearly antidepressants intake of at least 90DDD, of men and women who did not consume any antidepressants in 2008 (at the beginning of the observation period), according to their mobility status at the moment of their separation. The moving status is attributed from the moment of the mobility.

**Figure A6.**
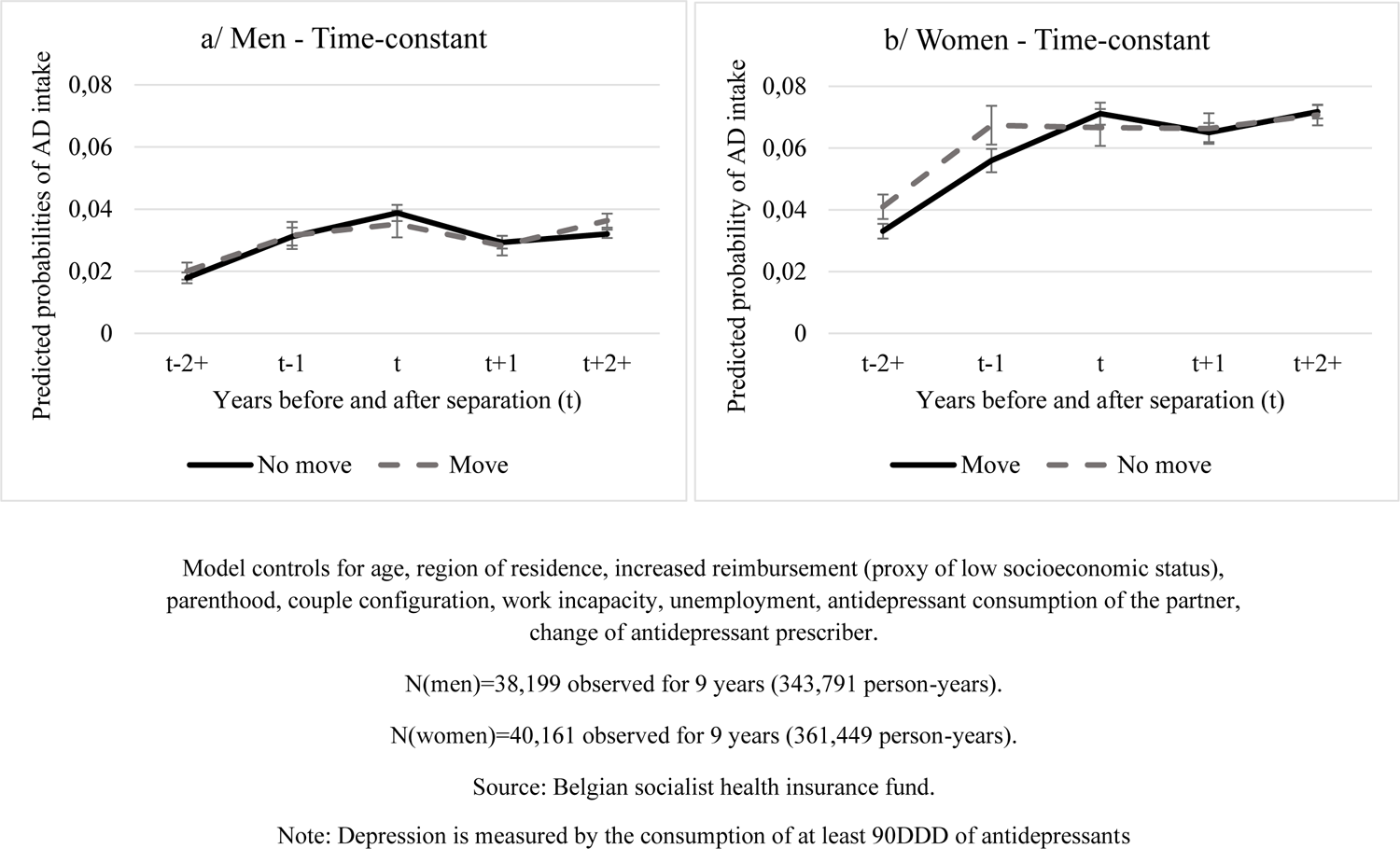
Predicted probabilities of a yearly antidepressants intake of at least 90DDD, of men and women who did not consume any antidepressants in 2008 (at the beginning of the observation period), according to their mobility status at the moment of their separation. The moving status is attributed during the whole period, before and after the mobility.

## ADDITIONAL MATERIAL

**Table A1.**
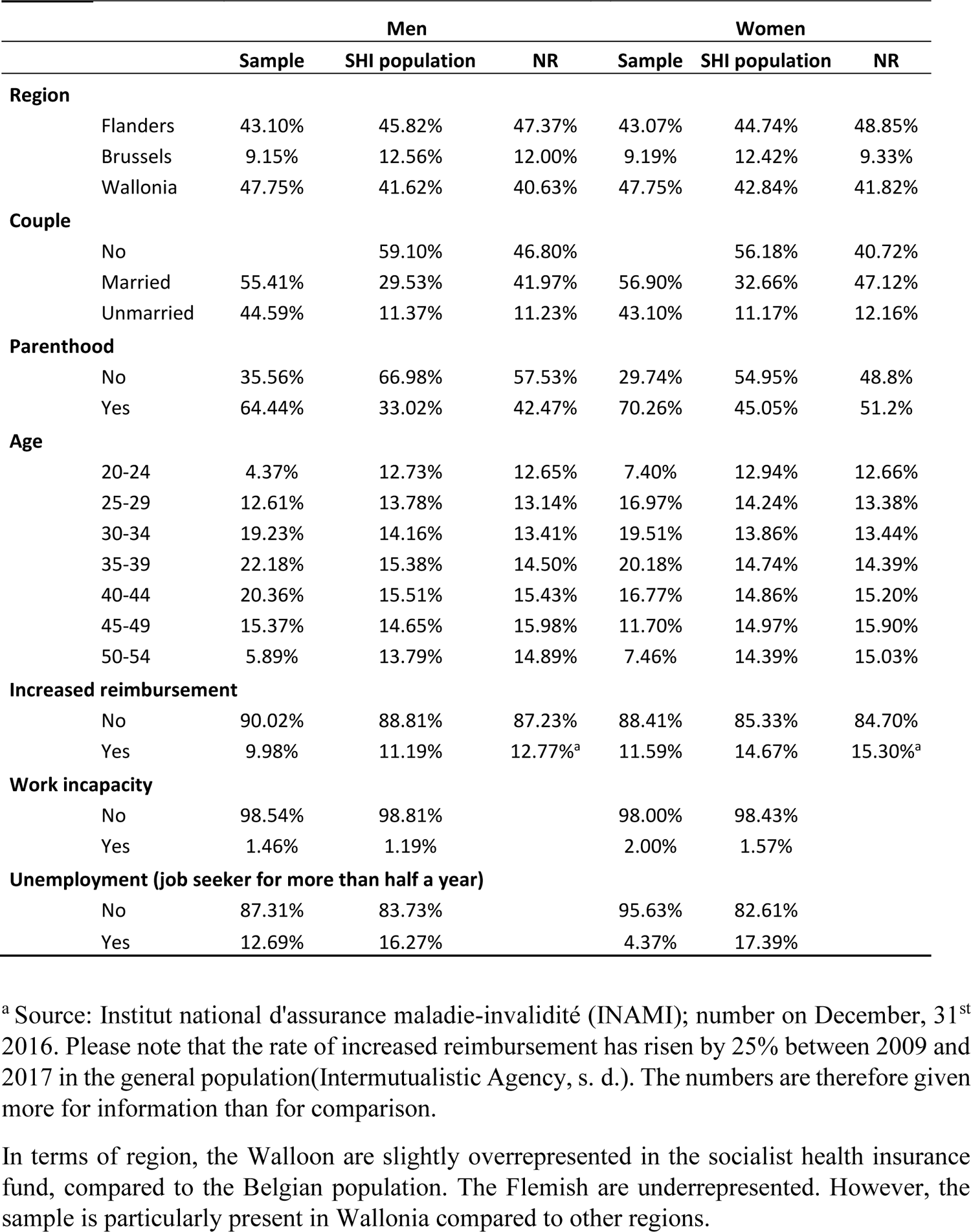
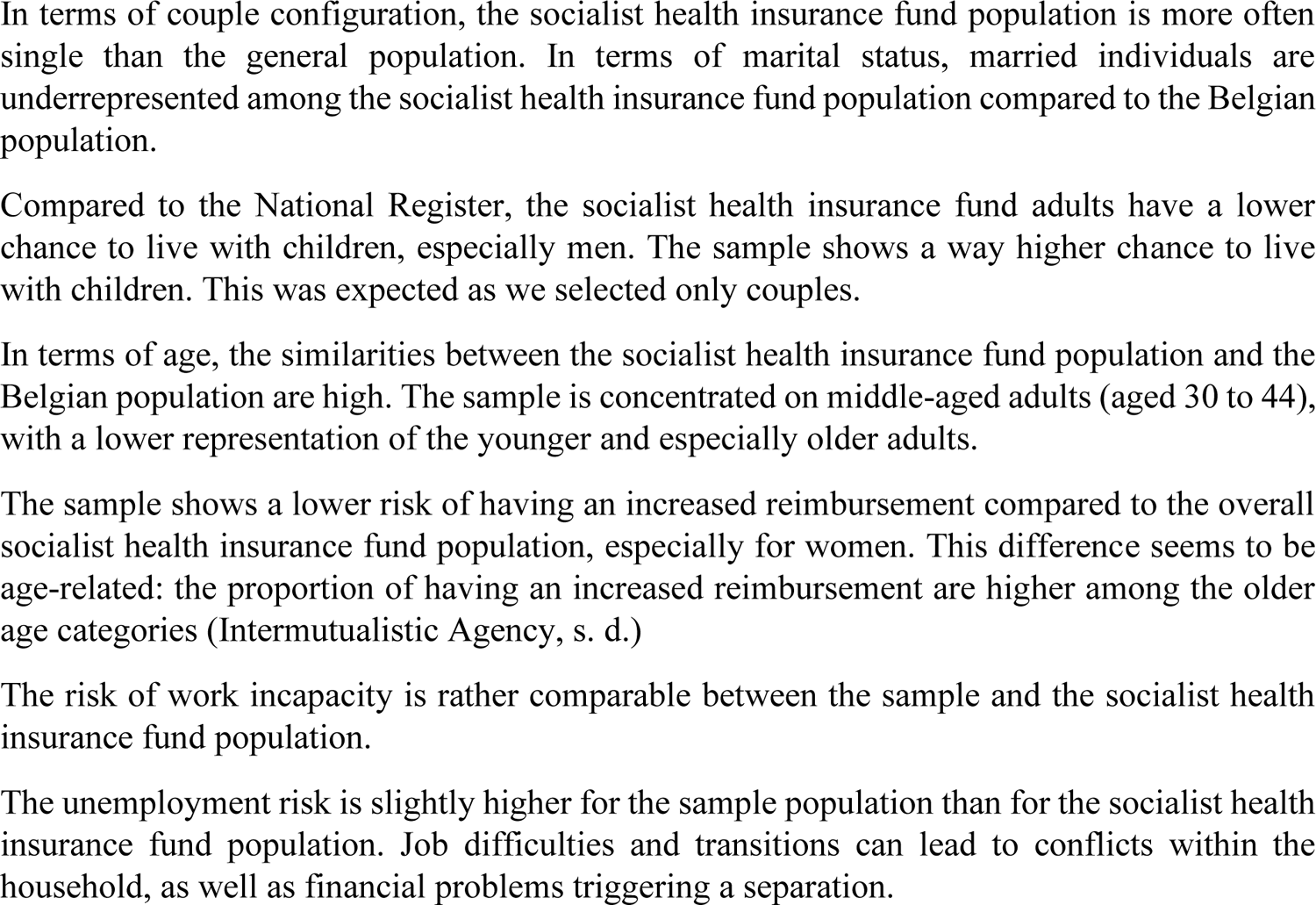
Characteristics of the sample population (adults aged 20 to 54 in 2009 and in a relationship in 2009 who will separate in the period 2009-2018), the socialist health insurance (SHI) fund population (all adults aged 20 to 54 in 2009 affiliated to the socialist health insurance fund in 2009) and the Belgian registered population (all adults aged 20 to 54 who are present in the National Register (NR) in 2009). Source for the Belgian population: Statistics Belgium and INAMI (National Institute for Health and Invalidity Insurances) for the increased reimbursement.

## Footnotes

1 https://www.solidaris.be/

2 Number of men and women differ because of the age restriction.

3 Nonetheless, being in a relationship does not mean a systematic lower consumption of mental health medication. It can also increase the individual’s antidepressants intake in two ways: by giving healthier habits, such as seeking for help in case of physical and/or mental issues, which can lead one’s access to mental health medication (Dupre & Meadows, 2007); by dedramatizing the consumption of such medicine when the partner already relies on such medicine.

